# Bayesian Ordered Lattice Design For Phase I Clinical Trials

**DOI:** 10.1101/2025.03.23.25324471

**Authors:** Gi-Ming Wang, Curtis Tatsuoka

## Abstract

We develop a new framework specifically for early Phase I clinical trials called Bayesian Ordered Lattice Design (BOLD). This study is motivated by two key factors. First, Phase I clinical trials typically involve relatively small sample sizes, which can make the use of prior information on dose-limiting toxicity (DLT) rates highly significant. To address this challenge, the proposed Bayesian methodology incorporates prior information and posterior updating to guide dose selection, toxicity monitoring, early stopping and identification of the maximum tolerable dose (MTD). Second, a natural ordering among toxicity probabilities across different dose levels can be utilized, with the idea being that analysis of dose-level posterior probabilities can and should acquire insights from data obtained at other dose levels, by leveraging their order relationship. Our proposed approach employs straightforward dose-level Bayesian specifications and relies on intuitive and clinically interpretable DLT rate posterior probabilities for decision making. Importantly, we show that it can often outperform popular methods, in terms of accuracy in determining the MTD. This Bayesian approach is also computationally simple and avoids simulation.

## 1. Introduction

Phase I clinical trials are designed to determine the maximum tolerable dose (MTD), which is the dose that has a probability of dose-limiting toxicity (DLT) closest to a predetermined target rate.^1^ Designs are comprised of two primary elements. The first element involves a set of criteria for making decisions regarding the allocation of dose level to each participant. The second element entails methods for determining when to stop and the identification of an MTD at the end of the trial.^2^ The process of identifying the appropriate dose for targeted therapies presents several challenges, including the need to enhance the accuracy of dose-finding efforts in order to reduce both human and financial costs, as well as to shorten development timelines.^3,4^ From both practical and ethical perspectives, it is crucial to minimize decision-making errors and reduce as possible the risk of exposing patients to sub-therapeutic or excessively toxic doses.^1^

A variety of Phase I trial designs have been established to determine MTD for single-agent studies. These designs can be categorized into three distinct types based on their statistical principles and methods of execution: algorithm-based, model-based, and model-assisted designs.^5^ The traditional 3+3 design is the most widely utilized algorithm-based approach in clinical trials, employing a straightforward set of predetermined rules to guide dose escalation and de-escalation.^6^ This design continues to be a predominant choice for Phase I trials, primarily due to its ease of use.^7^ Nevertheless, the 3+3 design has faced significant criticism for its inadequate operating characteristics,^8^ and its limited effectiveness in accurately identifying MTD and estimating DLT rates. Its heightened likelihood of administering sub-therapeutic doses to patients has also been noted.^9^

Model-based designs utilize a statistical model, such as a logistic model, to characterize the dose-toxicity relationship and facilitate dose adjustments.^8^ A prominent example of this design is the Continuous Reassessment Method (CRM).^8^ As new data is collected, the CRM method iteratively refines the model estimates after each cohort, aiding in the determination of appropriate dose selection.^9^ Compared to the traditional 3+3 design, CRM demonstrates superior accuracy in identifying and assigning a greater number of patients to MTD.^10^ However, it is important to note that the implementation of CRM can involve inherent statistical and computational complexities, such as in the estimation of the model.^9^

Model-assisted designs were created to integrate the benefits of both algorithm-based and model-based designs. Like model-based designs, model-assisted designs utilize a statistical model (e.g., the binomial model to estimate DLT rate) to aid in formulating decision rules. In a manner akin to algorithm-based designs, model-assisted designs can outline the rules for dose escalation and de-escalation, thereby simplifying implementation.^6^ Common examples of model-assisted designs include TPI (toxicity probability interval design), mTPI (modified toxicity probability interval design),^11^ Keyboard design (specifically, mTPI-2),^12^ and Bayesian Optimal Interval Design (BOIN).^7^ A notable characteristic of BOIN, one of the designs with which we will make a comparison, is that empirical proportions serve as the basis for dose selection. Its boundaries for escalation/de-escalation are derived from a Bayesian 3-class problem posed at each dose level, but Bayesian posteriors for the specific DLT rate values play no explicit role in dose selection. Instead, boundaries for the empirical proportion values determine dose selection decisions.

We propose a Bayesian framework aiming to enhance the accuracy in determining the MTD, a challenging issue in dose escalation trials. The primary motivation behind the proposed design is that Phase I clinical trials typically involve relatively small sample sizes, making prior information regarding dose toxicity highly significant. In clinical trials, the majority of conventional (frequentist) statistical techniques typically incorporate data from prior studies solely during the design phase, rather than as integral to the formal analysis.^13^ By utilizing prior information in a Bayesian framework, the accuracy of dose selection can be enhanced.

Furthermore, since a natural ordering of toxicity rates emerges across different dose levels, this can be recognized and utilized to enhance the precision of determining MTD. The underlying premise is that higher dose levels are anticipated to have higher DLT rates.^14^ The associated order constraints may be explicitly recognized through the posterior probabilities related to DLT rates across different doses.^15^ This linkage of posterior probabilities to reflect the natural order among dose DLT values allows for “learning” from response data across dose levels.

The objective of this research is to create a Bayesian Ordered Lattice Design (BOLD) that integrates prior knowledge and ordering constraints with empirical toxicity data for the purposes of adaptive dose selection, early stopping and classification of MTD in Phase I clinical trials. For the proposed BOLD, the incorporation of even weakly informative prior knowledge regarding dose toxicity in an order-constrained Bayesian manner can improve accuracy in scenarios where sample sizes are small.

The term “lattice” originates from the idea that the dose levels lie within a formal order structure. Dose levels in a single-drug trial can be perceived as a linearly ordered lattice. Web Figure 1 of the Supplementary Material illustrates the lattice diagram featuring five dose levels, which also includes top and bottom states. The bottom state signifies that an MTD is lower than the minimum dose. The top state signifies that the MTD is higher than the maximum dose. Classification involves selecting a state within the lattice which indicates the dose level believed to be the MTD. Related work on Bayesian sequential classification on lattice models includes Tatsuoka and Ferguson (2003),^16^ Tatsuoka (2014),^17^ and Tatsuoka, Chen and Lu (2022).^18^

## 2. Methodology

### Notation and Terminology

Suppose *J >* 1 dose levels. Denote DLT rate for dose *j* as *π_j_*, 1 ≤ *j* ≤ *J*. We assume the natural order constraints *π_j_* ≤ *π_k_*if *j < k*. Terminology from lattice theory that we use includes the notion of a *cover* and *anti-cover* of a dose level. Note that the cover for dose *j* is dose *j* + 1*, j < J*, and the anti-cover is dose *j* − 1*, j >* 1. These are the least upper bound and greatest lower bounds of dose *j* in the lattice. Let *φ* be the target DLT rate. *N_max_*is the maximum number of overall patients in the trial before stopping is invoked; 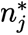 is the upper limit of patients at dose *j* before stopping the trial is considered; *n_c_*is the number of patients per cohort at a given stage, where a *stage* is defined as the period of administration and observation for DLTs after a dose has been selected; *n_j_* represents the total number of patients who have received dose level *j*, and *x_j_* denotes the number of those patients with observed DLTs, with 0 ≤ *x_j_* ≤ *n_j_*. Toxicity thresholds for dose *j* are denoted by *γ_j_*, in that if the posterior probability of *π_j_ > φ* exceeds *γ_j_*, then the administration for all doses *k*, *j* ≤ *k*, will no longer be considered. Finally, *τ* is a parameter reflecting a target probability value used in dose selection.

### Prior and conjugate posterior distribution

The implementation of BOLD requires assigning dose-level prior distributions for the respective DLT rates. We employ Beta distributions, which are represented as *π_j_* ∼ *Beta*(*α_j_, β_j_*) for dose level *j*. Importantly, Beta distributions have conjugate posteriors *π_j_*|*x_j_, n_j_* ∼ *Beta*(*α_j_* + *x_j_, β_j_*+ *n_j_* − *x_j_*). Bayesian updating is conducted after each stage.

In prior specification, we adopt the following guiding principles: 1) It is assumed that the prior distributions of respective DLT rates reflect the order of the dose levels, such as in terms of the prior means; 2) Priors should not be excessively informative, in that each dose must have the potential to be identified as MTD, even when only limited sample sizes are available. This principle supports equipoise. Specifically, this requires that the variances of the prior DLT rates should not be too small. 3) The prior means should also be within reasonable proximity to the target rate *φ*.

In the case of trials assuming non-informative priors, we define the parameters of the prior distribution *α* and *β* by setting the prior mean equal to the target rate, while the prior effective sample size (PESS), *α* + *β*, is suggested to match the standard cohort size at a given stage of administration. This will lead to a relatively weak prior in the sense that PESS is equal to one stage of dose administrations. It is interesting to note that if we were to assume a prior is a uniform distribution *Beta*(1, 1), the prior mean would be 0.5, which exceeds standard target toxicity values, although PESS = 2, which is weak. For informative priors, we will for instance specify the PESS of the dose considered most likely to be the MTD to be larger than other doses, which is equivalent to assigning a smaller prior variance.

### Order constraints on posterior probabilities

A key idea of BOLD is to impose order constraints to “share” updated information between dose levels after each stage. Another important feature of BOLD is that inferences for dose selection, toxicity monitoring, and early stopping rely on the same statistical basis, which we refer to as the Conjugate Posterior Probability of the DLT rate being Above the Target (CPAT), expressed as

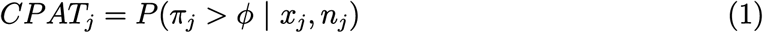

*CPAT_j_* represents the posterior probability that dose *j* is too toxic. Note that this posterior probability value provides the most direct and up-to-date insight on the most pertinent clinical question: Is the dose’s DLT rate safe or not, relative to the target *φ*? Larger values of (1) indicate that the DLT rate is more likely to be unacceptably high.

Order constraints on CPAT values can be implemented after each stage using the Pool-Adjacent-Violators Algorithm (PAVA),^19^ an isotonic regression method.^20,21^ PAVA substitutes any neighboring values that violate the non-decreasing ordering with a weighted average, with the number of treated patients as a weighting factor. This ensures that the resulting estimates exhibit monotonicity. We apply PAVA in a “local” manner, in the sense that only the CPATs of the current dose and its cover and anti-cover doses are included. This minimizes the constraints to those relevant for decision-making, since we only consider escalation/de-escalation in one dose level increments. Inclusion of doses in constraints with little or no observed data is thus minimized. A corresponding constrained *CPAT_j_* value following PAVA is referred to as the PAVA-adjusted Posterior Probability of the DLT rate being Above the Target (*PPAT_j_*). PPAT values are the basis for dose selection.

### Dose selection

BOLD jointly considers multiple dose levels in selection decisions based on “local” PPAT values. We restrict dose selection to either escalation, de-escalation, or maintaining the current dose. The proposed selection criterion involves identifying the dose for which the PPAT is nearest to *τ* among the current dose and its cover/anti-cover doses. If we denote this current set of dose levels as *J*^∗^, then dose *j*^∗^ ∈ *J*^∗^ is selected if it minimizes (2), where 0 *< τ* ≤ 0.5 is the PPAT threshold parameter:

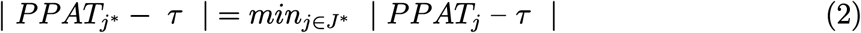

The default value of *τ* is 0.5. A PPAT value of 0.5 reflects the highest level of uncertainty as to the safety of that dose level (akin to a coin flip). Following observation from this stage, no matter the responses, we should gain improved insights one way or another as to whether the dose DLT rate is above or below the target.

When there is a tie in minimizing (2), with | *PPAT_j_*− *τ* | = | *PPAT_k_* − *τ* |*, j* ≤ *k*, the decision rule is as follows:

Case 1: If both PPAT values are below *τ*, choose dose *k*.

Case 2: If both PPAT values exceed *τ*, choose *j*.

Case 3: If *PPAT_j_* ≤ *τ* while *PPAT_k_ > τ*, choose dose *j*.

### Toxicity control

During experimentation, we will dynamically remove doses that are considered excessively toxic. Given a threshold *γ_j_ >* 0, a dose *j* is excessively toxic if *CPAT_j_ > γ_j_*. Note that if dose *j* is deemed too toxic, all doses *k*, *j* ≤ *k* ≤ *J*, are removed from further consideration. Furthermore, each dose can have its own threshold.

### Overdose control

Note that if *τ* in (2) is reduced from the default value of 0.5, this discourages escalation, as it is more attractive to select dose levels that have a lower probability that the DLT rate exceeds target than 0.5; in other words, a higher probability of being “safe”. Hence, overdose control can be specified intuitively and simply by adjusting the PPAT threshold parameter *τ*.

### Stopping rule

Stopping of the trial is invoked when:

Case 1: The minimum dose level (i.e., dose 1) is excessively toxic, as indicated by *CPAT*_1_ *> γ*_1_, suggesting that a MTD is lower than this minimum dose level.

Case 2: The number of patients receiving treatment in the trial reaches *N_max_*.

Case 3: The number of patients at dose *j* has reached 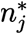 and dose *j* is the next stage selection.

### MTD identification

The MTD will be determined upon stopping. MTD identification relies on isotonically-regressed posterior means of DLT rates. As noted, if the lowest dose is found to be excessively toxic (*CPAT*_1_ *> γ*_1_), all doses are considered too toxic and no MTD is selected. Otherwise, we implement PAVA “locally” on the conjugate posterior means of the final selected dose, along with its cover and/or anti-cover doses and regard these doses as the potential MTD candidates. We have found that this can improve accuracy versus considering all posterior means after isotonic regression, which involves imposing more order constraints. Candidates for MTD must also have observed trial data. Let *J*^∗^ be the collection of candidates, and 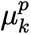 be the PAVA-adjusted posterior mean for dose *k* ∈ *J*^∗^. The dose *k* with smallest PAVA-adjusted posterior mean difference with the target DLT rate,

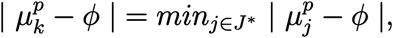

is selected as MTD. If a tie occurs among 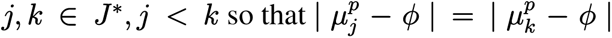, identification is as follows:

Case 1: If 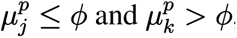, then dose *j* is selected as MTD.

Case 2: If 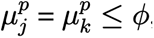, then dose *k* is selected.

Case 3: If 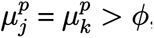, then dose *j* is selected.

### Design Inputs

In sum, the following are the design inputs: 1) the target DLT rate *φ*; 2) dose-level prior Beta distribution parameters for DLT rates for each *π_j_*; 3) *N_max_*, 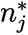 for each dose *j*, and *n_c_*; 4) toxicity thresholds *γ_j_* for each dose *j*; 5) PPAT threshold parameter *τ*. We assume that the initial dose level being administered is the lowest one, but this can also be specified.

### Flowchart of BOLD

The flowchart that outlines the procedures and guidelines for dose selection is presented in Figure 1. A summarized description of the flowchart is as follows: 1) Specify the target DLT rate *φ* and other inputs. 2) Assess prior information, determining whether the prior distribution will be informative or non-informative, and subsequently assign the Beta prior parameters that determine prior mean and variance, as well as toxicity threshold for each dose level. 3) Initiate the trial at the lowest dose. 4) Administer the dose level to *n_c_* patients and observe for DLTs. 5) Update the CPAT values, which represent the posterior probability that a dose is overly toxic. 6) Assess toxicity control based on *CPAT*_1_ exceeding *γ*_1_ or not, and consider early stopping of the trial. 7) Otherwise, continue monitoring if the overall sample size ceiling, *N_max_*, is reached. 8) Consider dose elimination of the currently administered dose *j* along with all the higher doses ≥ *j* by assessing the CPAT value of the current dose using its corresponding threshold *γ_j_*, and proceed to de-escalate the dose. 9) Otherwise, implement order constraints on the updated CPAT values to obtain PPAT values locally (current and cover/anti-cover dose(s)). 10) Select the next dose to either escalate, de-escalate or maintain the current dose based on minimizing the PPAT criterion in equation (2). 11) Monitor the current dose *j* for early stopping of the trial if the number of patients administered at dose *j* is at least 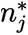 and if the decision in Step 10 is to maintain the current dose. 12) Repeat another stage of experimentation from Steps 4 to 11 until the trial is stopped. 13) Once stopping occurs, determine the MTD by considering PAVA-adjusted posterior mean DLT rates.

**Figure 1:**
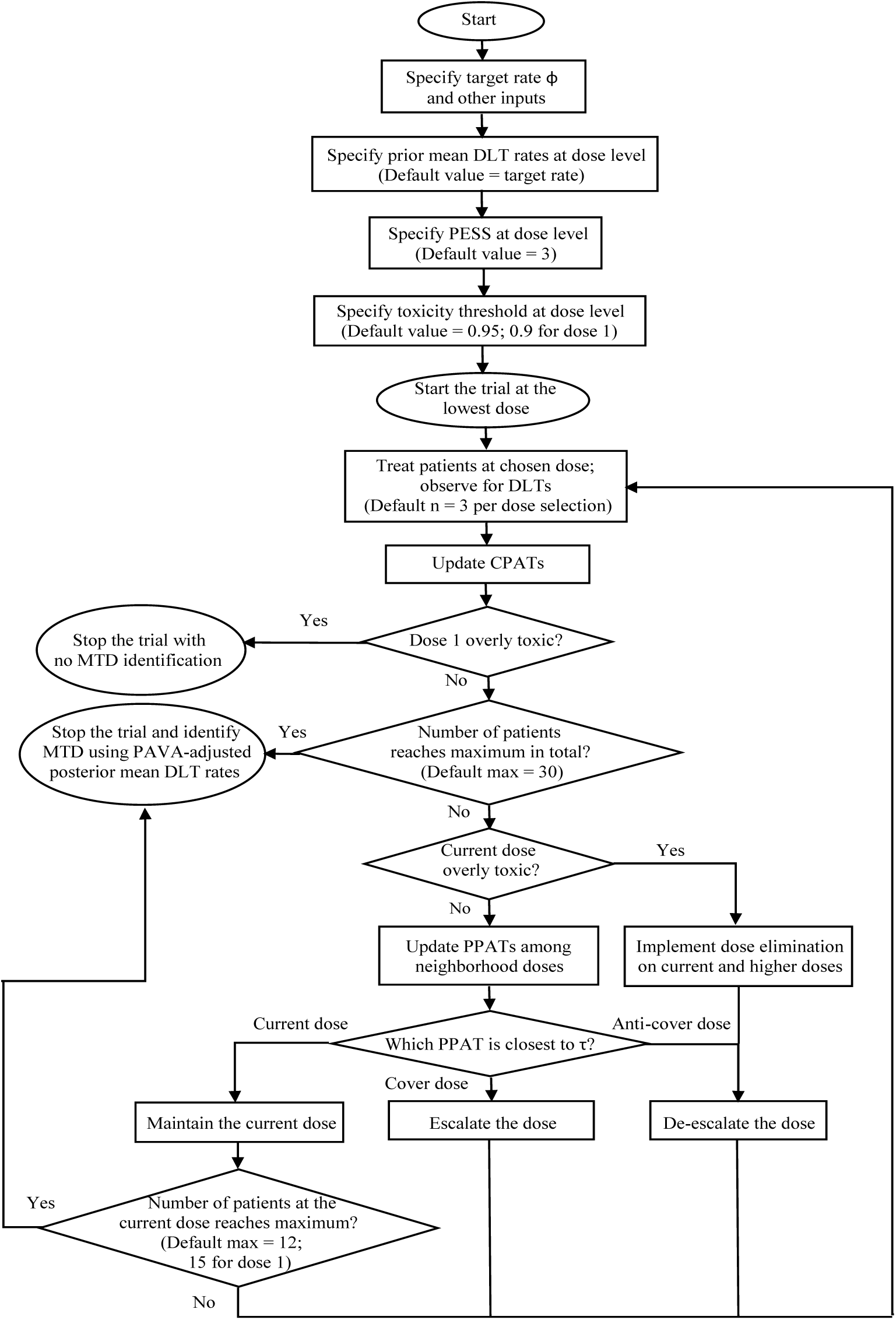
Flowchart of BOLD design for single-drug Phase I clinical trials.

## 3. Simulation

### Design and demonstration

The performance of BOLD for Phase I clinical trials can be evaluated via simulations conducted with R (code available at https://github.com/hiddenmanna1996/BOLD). In general, our default prior distribution, *Beta*(*α*, *β*), is set for all doses to have prior mean equal to the target rate *φ*, which reflects equipoise, and PESS (i.e. *α* + *β*) = 3, the typical standard cohort size (*n_c_*), which reflects a weak prior. For *φ* = 0.2, this is equivalent to *Beta*(0.6, 2.4); when *φ* = 0.25, it is *Beta*(0.75, 2.25). The upper limit of patient accrual during a trial (i.e., *N_max_*) is set at either 21 or 30. We focus on these smaller sample sizes since in our practice, timelines and duration are very much a concern for these trials, as well as the strong interest in moving forward to a dose expansion phase. Stopping occurs when the number of patients receiving any dose *j* reaches 12 (i.e. 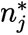 = 12) and concurrently dose *j* also is the next-stage dose selection decision. For BOLD, an exception is made for the lowest dose, where 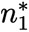 is set to 15 in order to enhance classification to the bottom state, which reflects that all doses are too toxic. Toxicity thresholds are set to *γ_j_* = 0.95, *j >* 1. For the lowest dose, *γ*_1_ is set to 0.9, again to enhance classification to the bottom state. Note that the bottom state is only classified to indirectly, since it is not an actual dose, and its classification is conducted by the toxicity threshold being exceeded for the lowest dose. Lastly, we set PPAT threshold parameter *τ* = 0.5, the default value.

As an illustration, assume that a trial considers 5 distinct dose levels, and the target response rate *φ* is set at 0.25. The MTD is assumed to be dose level 4. The prior mean DLT rate is assumed to be 0.25, the same as the target rate, for all the doses. Also assume the true DLT rates to be 0.1, 0.11, 0.12, 0.25 and 0.5 for doses 1 through 5, respectively. The updating of PPAT values after hypothetical toxicity responses and the corresponding dose selections are presented in Table 1. Note the preservation of order among the PPAT values. The final step of MTD identification is also illustrated with the final PAVA-adjusted posterior means.

**Table 1:** Demonstration of dose selections for a 5-dose model with non-informative prior, target rate *φ* = 0.25, and maximum total sample size *N_max_* = 30, whereas the process of dose selection utilizing PPAT involves escalation (E), de-escalation (D), or maintaining the current dose (M).

| Stage | Current dose level | Observed DLTs | Accumulated DLTs | Sample size of current dose | PPAT |  |  |  |  | Decision |
| --- | --- | --- | --- | --- | --- | --- | --- | --- | --- | --- |
|  |  |  |  |  | Dose 1 | Dose 2 | Dose 3 | Dose 4 | Dose 5 |  |
| 1 | 1 | 1 | 1 | 3 | 0.538 | 0.538 | -- | -- | -- | M |
| 2 | 1 | 0 | 1 | 6 | 0.289 | 0.410 | -- | -- | -- | E |
| 3 | 2 | 0 | 0 | 3 | 0.243 | 0.243 | 0.410 | -- | -- | E |
| 4 | 3 | 0 | 0 | 3 | -- | 0.152 | 0.152 | 0.410 | -- | E |
| 5 | 4 | 0 | 0 | 3 | -- | -- | 0.152 | 0.152 | 0.410 | E |
| 6 | 5 | 2 | 2 | 3 | -- | -- | -- | 0.152 | 0.850 | D |
| 7 | 4 | 1 | 1 | 6 | -- | -- | 0.152 | 0.289 | 0.850 | M |
| 8 | 4 | 0 | 1 | 9 | -- | -- | 0.148 | 0.148 | 0.850 | E |
| 9 | 5 | 3 | 5 | 6 | -- | -- | -- | 0.146 | 0.993 | D |
| 10 | 4 | 2 | 3 | 12 | -- | -- | 0.152 | 0.460 | 0.993 | M |
| PAVA-adjusted posterior mean DLT rate |  |  |  |  | -- | -- | 0.125 | 0.250 | 0.639 | Dose 4 is identified as MTD |

### Comparative analysis for non-informative prior

We begin a comparative analysis versus BOIN and CRM designs through simulations using non-informative priors. We first consider 5 doses, so that the possible MTD selections encompass “*<*1”, 1, 2, 3, 4, 5, and “*>*5”, where “*<*1” indicates that all doses are too toxic and “*>*5” signifies that all doses have DLT rates below the target. The simulations are carried out based on random scenarios of true DLT rates.^6^ In situations when the MTD corresponds to a dose level *j <* 5, the difference in DLT rates between the *j*-th value and the (*j* + 1)-th value, referred to as *upper δ*, is fixed at values of either 0.1, 0.15, 0.2, or 0.25. We carefully stratify DLT rate scenarios by these upper *δ* values, in order to carefully demarcate when performance differences between methods clearly emerge. Larger upper *δ* values reflect greater discriminability. Per scenario, 5 distinct DLT rates are generated and organized in ascending order within the range of (0.01, 0.7), imposing that the *j*-th value is the target rate *φ* and the (*j* + 1)-th value is equal to *φ* + upper *δ*. If *j >* 1, the difference in DLT rate between the *j*-th value and the (*j* − 1)-th value is restricted to be at least 0.05. Under these circumstances, the non-MTD DLT rate values are selected randomly from a uniform distribution and arranged in ascending order to correspond to the linear order among dose levels. In scenarios where all dose levels have DLT rates above MTD target, we randomly select 5 unique DLT rate values that are organized in ascending order within the range between 1.4 times the target rate and 0.7. In scenarios where all dose level rates are below MTD target, we randomly select 5 unique values organized in ascending order between 0.01 to 0.6 times the target rate.

For every one of the 4 upper *δ* values (i.e., 0.10, 0.15, 0.20, and 0.25), a total of 50 random scenarios are generated for each of the 7 MTD levels (i.e., *<*1, 1, 2, 3, 4, 5, and *>*5), culminating in a grand total of 1,400 random scenarios. For each random scenario, 100 simulations of response sequences are generated. This number of random scenarios per dose level and the corresponding number of simulations per scenario are the same throughout all examples. The simulations for BOIN and CRM are conducted utilizing R functions get.oc (Generate operating characteristics for single agent trials) and bcrm (Bayesian continual reassessment method for Phase I dose-escalation trials), respectively. The dose-toxicity response curve associated with the CRM design is assumed to follow a one-parameter logistic model with uniform prior on the interval (0, 10).^22^ The prior probabilities of toxicity in the CRM design for each dose level (i.e., skeletons) are generated by the R function getprior() according to default specifications suggested by Lee and Cheung (2009).^23^ The value of *γ*_1_, the toxicity threshold for dose 1, is set at 0.9 for all the three designs (BOLD, BOIN and CRM). For dose level *j >* 1, *γ_j_* for BOLD and BOIN is established at 0.95 (the bcrm code currently does not offer this option). The value of 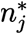 for all *j* is set to be 12 for BOLD, BOIN and CRM, except where 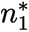 = 15 for BOLD as previously indicated. More attention is given to the BOLD versus BOIN comparison, due to their methodological similarity.

Accuracy is quantified as the proportion of trials that successfully identify the MTD, with the standard deviation of accuracy rate computed from the simulations across each of the scenarios. Efficiency is assessed by the mean and standard deviation of number of patients treated from across the scenarios. Figure 2 depicts the bar charts of accuracy by MTD levels with non-informative priors, stratified by *N_max_*and upper *δ*, with *φ* = 0.25. Bar charts illustrating efficiency (mean patient count and corresponding standard deviation) are presented in Web Figures 2 and 3.

**Figure 2:**
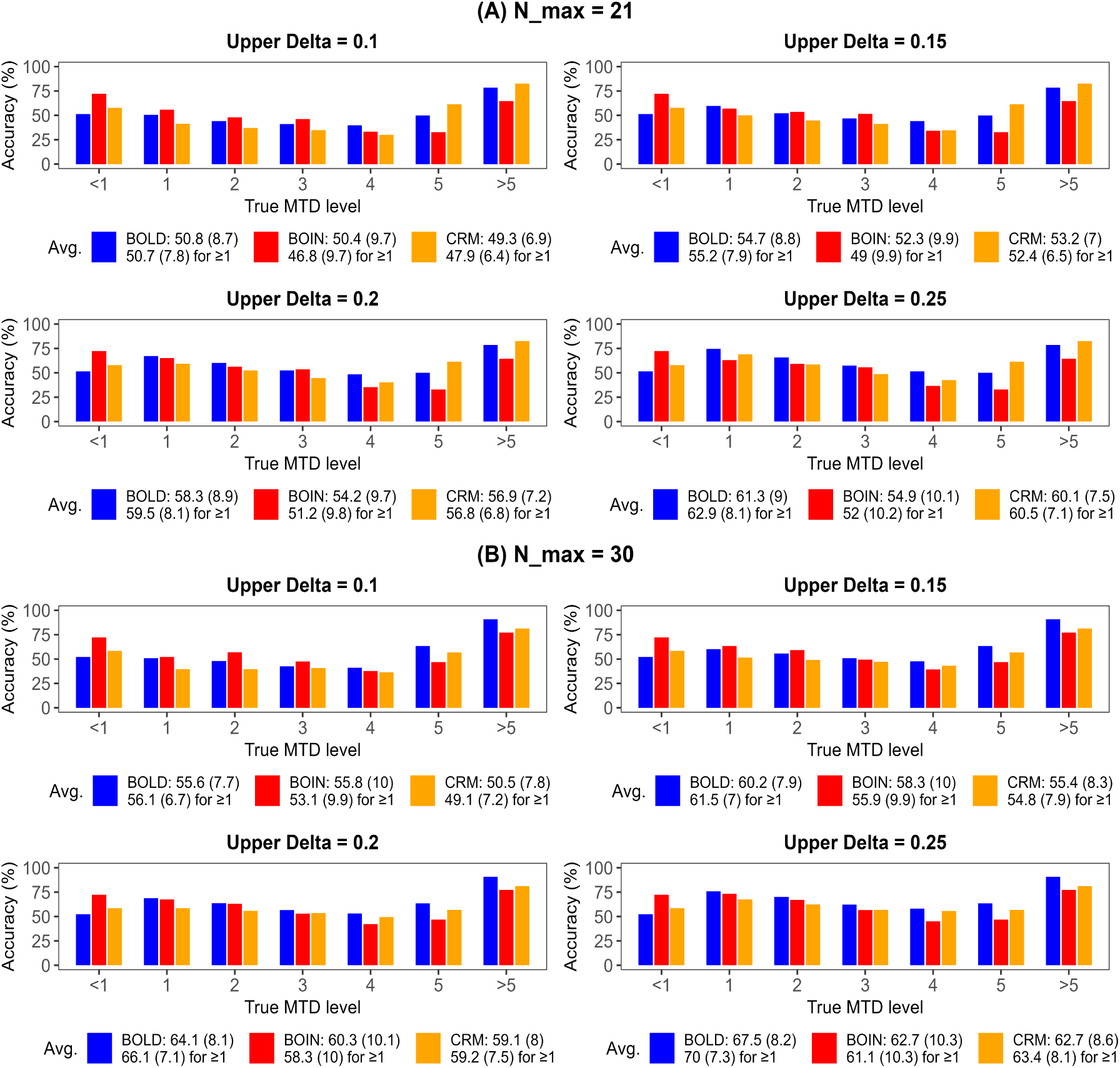
Accuracy of BOLD, BOIN, and CRM design, represented by the average of accurate MTD identifications (%) across scenarios, for 5-dose model with non-informative prior by MTD level and stratified by *N_max_* and upper *δ*, assuming random scenarios of true DLT rates, at target rate *φ* = 0.25. The legend denotes the average and SD (in parenthesis) for all the 7 MTD levels (1st row) and for the MTD levels that are *≥* 1 (2nd row).

In these figures, averages are reported based on all 7 scenarios, as well as for all scenarios except the case when all DLT rates exceed the target (the bottom state). The latter select set of scenarios would be of interest if an investigator was confident that at least one of the dose levels has a DLT rate at or below target. Hence, these average values are also of practical interest. Average accuracy rates for all designs at the upper *δ* value of 0.10 are around 50% or a bit higher, which underscores the limitations of small sample analysis when there is no clear discrimination between dose level DLT rates. Overall, these figures suggest that in many scenarios, the average accuracy across dose levels of BOLD exceeds BOIN and CRM designs. Specifically, at the dose level, this advantage becomes more pronounced when the MTD is at higher dose levels, and as the upper *δ* value increases. Moreover, the accuracy rates across dose levels seem to be more consistent for BOLD, in terms of maximizing the lowest of the dose-level accuracy rates (maximin property). Compared to BOIN, BOLD and CRM appear to leverage larger DLT rate differences more effectively as accuracy improvements are more pronounced. This is due to BOLD and CRM selecting dose across different levels at each stage of selection, as opposed to focusing on a specific dose until an escalation/de-escalation decision is made, as does BOIN. On the other hand, BOIN’s approach does lead to superior identification of the bottom state when it is true. These respective advantages hold at *N_max_* = 21 and 30. Note that for all these methods, accuracy improves as *N_max_* increases. Standard deviations of accuracy rates across scenarios are also tracked in Figure 2. CRM has smaller standard deviations of accuracy when *N_max_* = 21, but the differences decrease at *N_max_* = 30. Compared to BOIN, BOLD generally has smaller standard deviations in accuracy rates across simulations.

Figure 3A displays the plots of true DLT rates across dose levels from 50 random scenarios for each MTD level of a 5-dose model with upper *δ* = 0.15, MTD levels 1 to 5, *φ* = 0.25, and *N_max_* = 21. The plots also display accuracy comparisons between BOLD and BOIN for each scenario, demarcating *>*10% differences in mean accuracy (computed from the larger value) across 100 simulations within each scenario. Similar plots when upper *δ* = 0.2 are shown in Figure 3B. These plots indicate that BOLD’s relative advantage in accurate MTD identification increases at higher dose levels. The cases when the bottom (*<* 1) and top state (*>* 5) are true are not shown. However, as mentioned, BOIN has clear advantages when all dose levels are overly toxic for the bottom state. BOIN also seems to have an advantage specifically at the middle level doses in the scenarios when the lower dose levels have DLT rates closer to zero. This leads the algorithm to quickly escalate out of the lower dose levels, which can be an issue more generally for BOIN when higher doses are the MTD.

**Figure 3:**
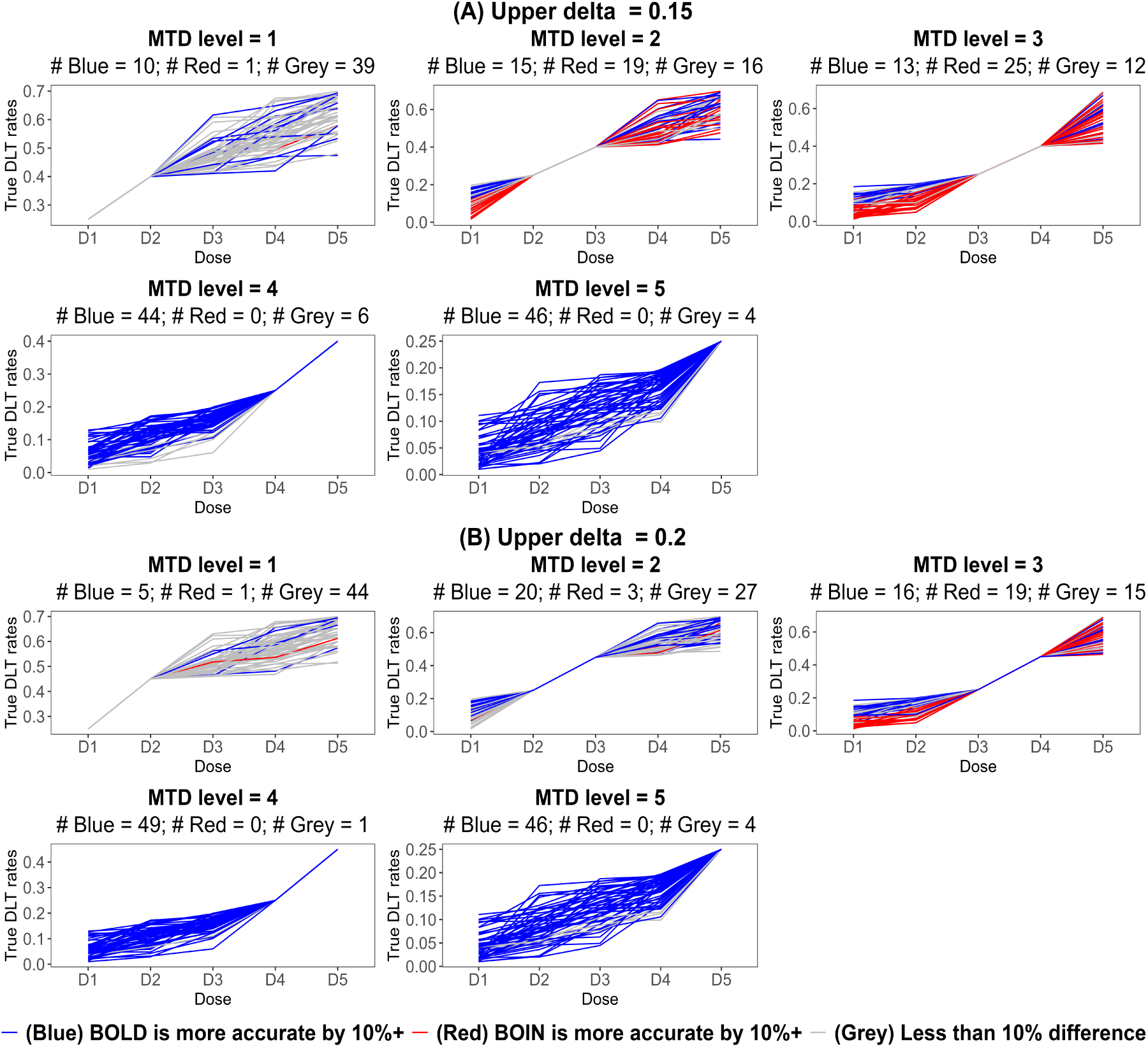
Plots of true DLT rates of a 5-dose model with 50 random scenarios for each MTD level, whereas upper *δ* = 0.15 and 0.2 at MTD levels 1 to 5, *φ* = 0.25, and *N_max_*= 21, alongside an accuracy comparison between BOLD and BOIN for each scenario, where the 10% difference of accuracy is defined as from the larger value. The plots for MTD = 5 are identical across both upper *δ* values. Although not displayed, there are zero blue and 47 reds for MTD *<* 1, and 43 blues and zero red for MTD *>* 5.

Efficiency is also considered. In Web Figure 2, CRM requires a smaller number of patients for *φ* = 0.25. We do note that when *N_max_* is larger, differences between BOIN and CRM in average number of patients have not arisen in other simulations.^1^ In other settings, there are no clear differences between BOLD and BOIN, and BOLD can have lower average patient counts when not including the bottom state. In terms of standard deviations in the number of patients across scenarios, as in Web Figure 3, BOLD generally has similar or smaller values than CRM, and BOIN clearly has the highest values. This is an indication of the consistency and stability of the designs in terms of patient numbers.

Web Figures 4 through 6 illustrate accuracy and Web Tables 1 and 2 present efficiency results at target *φ* = 0.15, 0.20 and 0.30 for *N_max_* = 21 and 30. Generally, patterns and trends in accuracy and efficiency appear similar as when *φ* = 0.25. In general, BOLD performs relatively poorly when the bottom state is true. For higher dose levels, BOIN performs relatively poorly for MTD levels 4 and 5, especially for the smaller study sample size when *N_max_* = 21. The selected CRM model can perform relatively poorly for the second highest dose level (dose *J* − 1). CRM also has a reduction in the average number of patients as *φ* increases, and BOLD often has smaller average standard deviations.

Web Table 3 shows evaluations of overdosing rates (the proportion of patients treated with doses higher than the actual MTD), underestimation rates (the proportion of trials with identified MTD being lower than the actual MTD), and overestimation rates (the proportion of trials with identified MTD being higher than the actual MTD) with non-informative priors and 5-dose lattice. The assumed *φ* values are 0.25 and 0.3, for *N_max_* = 21 and 30 and upper *δ* = 0.15. Note that when dose level 5 is the true MTD, overdosing and overestimation do not occur. Similarly, there is no possibility of underestimating when the bottom state is true and every dose is toxic. The table indicates that the overdosing and overestimation rates for BOLD are clearly higher than those for BOIN, while the underestimation rates of BOLD are comparatively lower. However, note that accuracy rates for the higher dose levels are much higher for BOLD, and that the maximin rates for BOLD are clearly higher as well. To some degree, relatively higher overdose rates are a necessary cost to more quickly investigate higher dose levels and to achieve more balanced accuracy performance across doses. This is especially important in smaller sample settings. There is no clear advantage for BOLD versus CRM in lower overdose rates.

To explore the impact of larger sample sizes, a sample of scenarios with design parameters as described in Figure 2 was simulated for *N_max_* = 36. Some adjustment in the 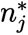-values was made to take advantage of the higher ceiling, so that 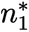 = 18, 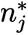 = 15*, j >* 1. Respective differences in accuracy, efficiency, and overdosing between the methods remain about the same as for the smaller sample cases. Relative advantage and disadvantages of BOLD versus BOIN should extend to larger sample sizes as well. Results are not presented here and we refer to the R app for the recreation of results.

Simulations are also conducted for 2, 3, and 4-dose lattices. Web Tables 4 and 5 illustrate the accuracy, efficiency, and overdosing rates of BOLD, BOIN and CRM with non-informative priors by lattice size for *N_max_*= 21 and 30 respectively, utilizing random scenarios of true DLT rates with upper *δ* = 0.15 and 0.20 at *φ* = 0.25. The overall average performance across methods is similar for all of these lattice sizes, although by the 4-dose lattice, an advantage for BOLD emerges at *N_max_* = 21. The reduction in the relative advantage for BOLD in overall average accuracy for fewer dose levels is due in part to averaging over a smaller number of scenarios that still include the bottom state, where BOLD performs relatively poorly. When only non-bottom state scenarios are considered, BOLD continues on average to outperform BOIN and CRM in terms of accuracy. Note that for all designs, there is an increase in the accuracy rate as the number of dose levels decreases, accompanied by a lower mean number of patients administered treatment before stopping is invoked.

Overdose control is explored in Web Table 6. As indicated in the Methodology section, adjusting the PPAT threshold parameter (*τ*) may assist in reducing the overdosing rate. A summary is reported for the accuracy, efficiency, and overdosing rates of BOLD at a 5-dose model for *φ* = 0.25 and 0.3 and for *N_max_* = 21 and 30. The *τ* values considered are 0.5 (default), 0.48, and 0.45. Random scenarios of true DLT rates are generated with upper *δ* = 0.15. The table indicates that for *φ* = 0.3, the overdosing rates with *τ* = 0.48 are significantly reduced in comparison to *τ* = 0.5. Additionally, it demonstrates that both the accuracy rates and patient counts experience only a slight decline with lower *τ* values for *φ* = 0.3. For *φ* = 0.25, this approach has less dramatic impact (see Discussion).

### Comparative analysis with a titration design

An important instance of Phase I clinical trial designs is the use of titration, which involves initially administering sub-therapeutic doses that are expected to be safe. This is prudent especially for first-in-human studies. If toxicity events arise with these first doses, the drug is perceived as potentially hazardous and it may be unwise to proceed further. Consequently, setting a lower toxicity threshold for more aggressive stopping and a lower prior toxicity rate to reflect the expected safety of a sub-therapeutic dose aligns with this principle.

To compare the performance of BOLD under titration, we select iBOIN (app) as the benchmark. The iBOIN design maintains the same decision-making framework as BOIN; however, it distinguishes itself by incorporating historical data into the dose selection boundaries, which can be tailored to specific doses. The historical information used by iBOIN regarding DLT rates is termed the “skeleton”,^6^ representing the anticipated DLT probability for each of the dose levels with a corresponding PESS value. Importantly, this step does not involve specifying associated prior distributions for dose-level DLT rate values and hence does not directly employ standard Bayesian updating.

Table 2 illustrates the simulated accuracy, efficiency, overdosing, percentage of trials reaching toxicity threshold at dose 1, and average number of treated patients at dose 1 for BOLD and iBOIN under a titration model involving 3, 4, or 5 doses, including a single sub-therapeutic dose, along with select values of *φ*, *N_max_*, and upper *δ*. The PPAT threshold parameter (*τ*) is set at 0.45 for overdose control, and *γ_j_* = 0.95, j *>* 1. The toxicity threshold *γ*_1_ for the sub-therapeutic dose is 0.6, reflecting a high sensitivity to safety concerns. Additionally, its prior mean toxicity rate for BOLD and the skeleton of iBOIN are both fixed at 0.05, while the true DLT rate in simulations for this lowest dose is restricted to be less than 0.05. Given that the sub-therapeutic dose is not considered as a possible MTD, results for the single sub-therapeutic dose are not included in the table. Instead, we use the notation “*<*2” to denote that the MTD is less than dose 2, where dose 2 is the lowest possible dose that can be identified as MTD for a next phase trial. The state “*<*2” is treated as the bottom state in this setting. This state can be classified to if: 1) dose 1 is found to be too toxic; 2) dose 2 is too toxic; 3) 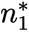 is reached and the next stage dose selection is dose 1, either through maintenance or by de-escalation from dose 2. Here, the value of 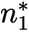 for BOLD is established at 9, as we want to limit the number of patients administered this dose, and 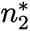 = 15, the value previously used for the first dose in non-titration cases. The iBOIN app does not allow for dose-level values of 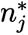, so the value is 12 for all doses.

**Table 2:**
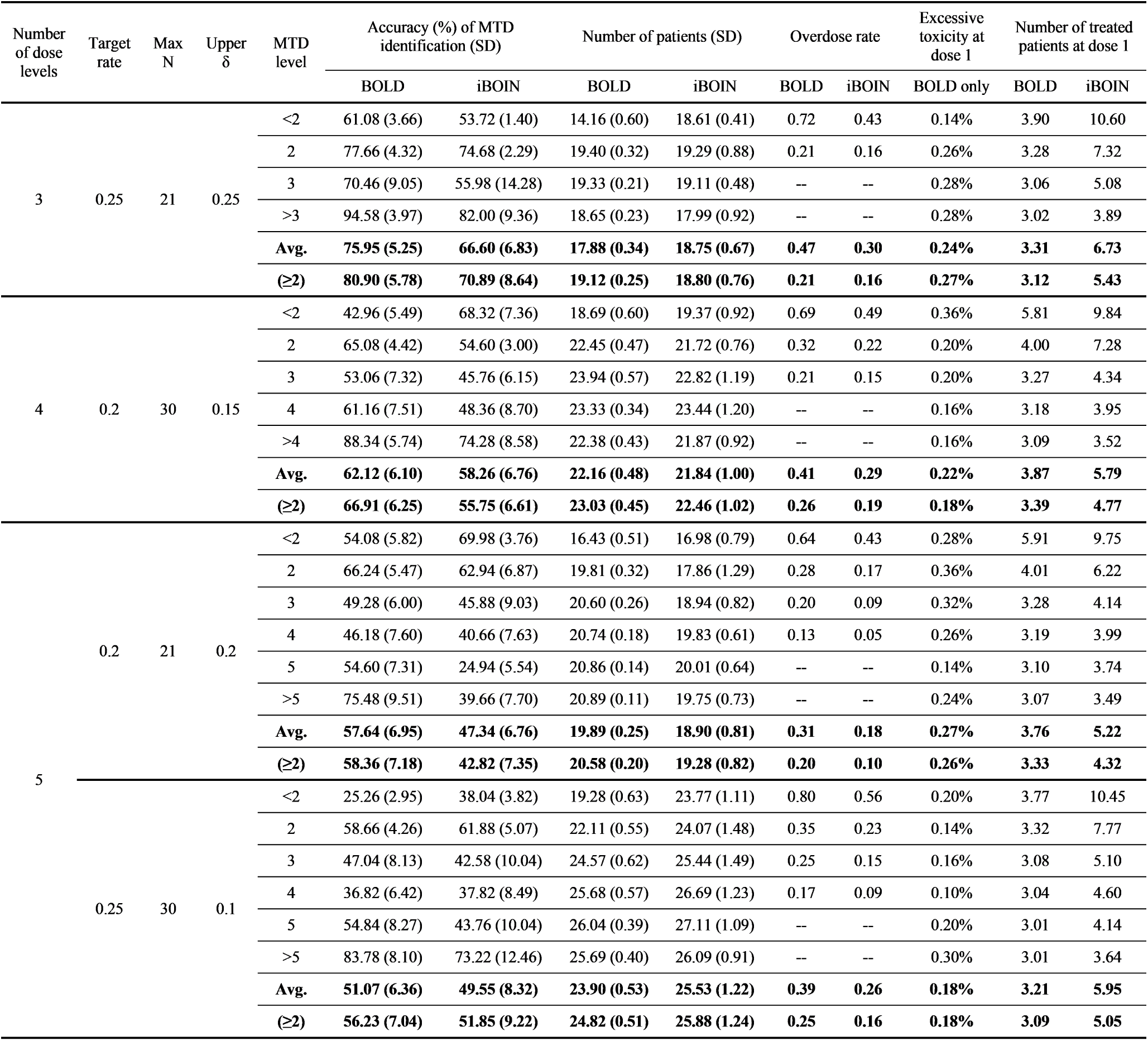
Summary of performance of BOLD and iBOIN under the titration assumption with sub-therapeutic dose 1, including accuracy, efficiency, overdose, percentage of trials reaching toxicity threshold at dose 1, and average number of treated patients at dose 1. Models involve 3, 4, or 5 doses, along with a selection of *φ*, *N_max_*, and upper *δ* of random scenarios, where *γ*_1_ = 0.6 and *γ_j_* = 0.95 for *j >* 1, prior mean toxicity rate for BOLD and the skeleton of iBOIN are both 0.05 for dose 1 and *φ* otherwise. The true DLT rate for dose 1 is below 0.05. PPAT threshold parameter *τ* is set to be 0.45 for BOLD. The state “*<*2” represents the bottom state and includes the cases in which the MTD level is “*<*1” or 1. “Avg.” denotes the average for all the MTD levels and “(*≥*2)” denotes the average for the MTD levels that are *≥* 2.

The table shows that for these design configurations, BOLD clearly surpasses iBOIN in terms of accuracy at the various dose levels (lattice sizes) in a titration design. This holds across a range of *φ*, *N_max_* and upper *δ* values, although for upper *δ* = 0.10 the advantage for BOLD is less. BOLD still generally does relatively worse in correctly identifying the bottom state. The average number of patients treated with the sub-therapeutic dose, and the percentage of times it is mistakenly classified as too toxic are also reported. For BOLD in particular, the impact of erroneous early stopping, as well as the number of patients receiving the sub-therapeutic dose, is small.

### Comparative analysis for informative priors

We also conduct a comparative analysis with iBOIN for trials with informative priors favoring a specific dose level. Two types of informative prior specifications that we consider are: 1) A favored dose is associated with a greater PESS value, such as 6 versus 3, in contrast to the other doses, while the prior means (skeletons) are all equal to *φ*. Note that larger PESS values indicate relatively smaller posterior variance and hence is more informative. 2) Assume that the prior means (skeletons) are mildly distinct, with the favored dose having the prior mean (skeleton) equal to *φ* while other prior means (skeletons) are within a close range (e.g., ±0.05 from *φ*), with values satisfying the linear order and PESS = 3 for all doses. Both approaches preserve equipoise and are only mildly informative. For each type of informative prior specifications, simulations for 5 dose levels are conducted. The settings for toxicity thresholds and random scenarios remain consistent with those employed for the trials with non-informative priors.

First, the MTD is assumed to correspond to the favored dose level, to assess how “correctly” specified informative priors enhance performance. Second, we also perform sensitivity analyses related to prior specifications. In trials with informative priors, specifications may incorrectly favor the wrong dose in real-world applications. We thus evaluate the robustness of the proposed priors under misspecification. Two possible situations are examined: either the anti-cover dose or the cover dose of the favored dose is the true MTD.

Table 3 summarizes the accuracy, efficiency, overdosing, underestimation, and overestimation rates of BOLD and iBOIN by averaging performance across randomly generated scenarios with informative priors for a 5-dose lattice, and with *φ* = 0.25, *N_max_* = 21 and 30, and the lower and higher range of upper *δ* values, 0.1 and 0.25. The table demonstrates that BOLD generally exhibits higher accuracy compared to iBOIN across scenarios, even when the MTD diverges from the favored dose. There is also greater gain in incorporating this information when the favored dose coincides as the MTD, as reflected by even greater advantages in accuracy favoring BOLD compared with non-informative settings. The mean patient counts for BOLD are often slightly lower than for iBOIN, and again the corresponding standard deviations of number of patients for BOLD are lower. Web Tables 7 and 8 present a summary of accuracy, efficiency, overdosing, underestimation, and overestimation rates of BOLD and iBOIN by MTD level using informative prior PESS specifications for *N_max_*= 21 and 30 respectively, with a 5-dose lattice, *φ* = 0.25, and upper *δ* = 0.1 and 0.25. Web Tables 9 and 10 present the summary of results from informative prior mean/skeletion specifications. For clarification, in these tables, if the true MTD is dose 1 while also being the anti-cover of the favored dose, then the favored dose is dose 2. Conversely, if the true MTD is dose 1 while also being the cover dose of the favored dose, then the favored dose is the bottom state. If the true MTD is dose 5 and it also is the anti-cover of the favored dose, then the favored dose is the top state; if it is the cover dose, then the favored dose is dose 4. Note that specification of informative prior means (skeletons) to favor a dose compared to specifying a higher PESS value (smaller prior variance) is more robust when the favored dose is not MTD, in that the cover and anti-cover dose accuracy rates when they are the MTD are higher.

**Table 3:** Summary of average accuracy, efficiency, overdose, underestimation, and overestimation rates of BOLD and iBOIN across MTD levels 1 to 5, utilizing informative priors with 5-dose lattice, *φ* = 0.25, *N_max_* = 21 and 30, and chosen upper *δ* values of 0.1 and 0.25 for random scenarios, with prior specifications (spec.) of PESS and prior mean. Either the favored dose is the MTD, the cover dose of the favored dose is MTD, or the anti-cover dose is MTD, whereas the cover dose for dose 5 corresponds to the top state in Web Figure 1 and the anti-cover dose for dose 1 to the bottom state.

| N<br>max | Prior<br>spec. | True<br>MTD | Upper $\delta$ | Accuracy (%) of MTD<br>identification (SD) | | Number of patients (SD) | | Overdose rate | | Underestimation<br>rate | | Overestimation<br>rate | |
| --- | --- | --- | --- | --- | --- | --- | --- | --- | --- | --- | --- | --- | --- |
|  |  |  |  | BOLD | iBOIN | BOLD | iBOIN | BOLD | iBOIN | BOLD | iBOIN | BOLD | iBOIN |
| 21 | PESS | Favored<br>dose | 0.1 | 51.76 (7.53) | 44.91 (8.54) | 19.99 (0.22) | 19.13 (0.51) | 0.27 | 0.19 | 0.24 | 0.39 | 0.31 | 0.20 |
|  |  |  | 0.25 | 66.16 (7.83) | 54.08 (9.03) | 19.89 (0.23) | 19.09 (0.53) | 0.21 | 0.16 | 0.24 | 0.39 | 0.13 | 0.09 |
|  |  | Anti-cover of<br>favored | 0.1 | 39.98 (6.50) | 39.15 (7.18) | 20.01 (0.24) | 19.13 (0.51) | 0.30 | 0.19 | 0.28 | 0.41 | 0.40 | 0.25 |
|  |  |  | 0.25 | 55.69 (7.47) | 47.59 (7.78) | 20.00 (0.24) | 19.09 (0.53) | 0.24 | 0.16 | 0.28 | 0.41 | 0.20 | 0.14 |
|  |  | Cover<br>dose of<br>favored | 0.1 | 43.26 (6.62) | 39.81 (7.30) | 20.08 (0.23) | 19.13 (0.51) | 0.28 | 0.19 | 0.30 | 0.43 | 0.33 | 0.21 |
|  |  |  | 0.25 | 57.60 (7.05) | 48.23 (7.81) | 19.99 (0.24) | 19.09 (0.53) | 0.21 | 0.16 | 0.31 | 0.44 | 0.14 | 0.10 |
|  | Prior<br>mean /<br>Skeleton | Favored<br>dose | 0.1 | 54.52 (6.68) | 45.93 (8.29) | 19.98 (0.19) | 19.12 (0.49) | 0.29 | 0.19 | 0.20 | 0.38 | 0.32 | 0.20 |
|  |  |  | 0.25 | 68.96 (6.48) | 54.53 (8.87) | 19.88 (0.20) | 19.09 (0.50) | 0.22 | 0.16 | 0.20 | 0.38 | 0.13 | 0.09 |
|  |  | Anti-cover of<br>favored | 0.1 | 46.69 (5.62) | 43.45 (7.47) | 20.14 (0.19) | 19.16 (0.44) | 0.39 | 0.20 | 0.17 | 0.36 | 0.46 | 0.25 |
|  |  |  | 0.25 | 65.43 (6.17) | 52.62 (8.42) | 20.07 (0.19) | 19.05 (0.46) | 0.30 | 0.17 | 0.18 | 0.36 | 0.21 | 0.14 |
|  |  | Cover<br>dose of<br>favored | 0.1 | 47.73 (6.91) | 41.40 (7.20) | 20.02 (0.21) | 19.11 (0.49) | 0.27 | 0.19 | 0.28 | 0.43 | 0.30 | 0.19 |
|  |  |  | 0.25 | 61.75 (7.15) | 49.97 (7.73) | 19.94 (0.21) | 19.07 (0.51) | 0.21 | 0.16 | 0.29 | 0.43 | 0.12 | 0.09 |
| 30 | PESS | Favored<br>dose | 0.1 | 55.97 (7.03) | 48.96 (9.35) | 24.26 (0.51) | 25.10 (0.89) | 0.33 | 0.21 | 0.20 | 0.36 | 0.30 | 0.19 |
|  |  |  | 0.25 | 71.92 (7.39) | 56.97 (9.86) | 23.67 (0.49) | 24.60 (0.92) | 0.23 | 0.17 | 0.20 | 0.38 | 0.10 | 0.07 |
|  |  | Anti-cover of<br>favored | 0.1 | 43.52 (6.85) | 44.46 (9.54) | 24.33 (0.52) | 25.10 (0.89) | 0.36 | 0.21 | 0.23 | 0.38 | 0.42 | 0.22 |
|  |  |  | 0.25 | 61.36 (7.41) | 52.42 (9.56) | 24.02 (0.47) | 24.60 (0.92) | 0.27 | 0.17 | 0.24 | 0.40 | 0.18 | 0.10 |
|  |  | Cover<br>dose of<br>favored | 0.1 | 46.73 (6.59) | 44.80 (9.04) | 24.51 (0.52) | 25.10 (0.89) | 0.33 | 0.21 | 0.27 | 0.39 | 0.33 | 0.20 |
|  |  |  | 0.25 | 63.34 (7.39) | 52.66 (9.14) | 23.97 (0.49) | 24.60 (0.92) | 0.24 | 0.17 | 0.27 | 0.41 | 0.12 | 0.08 |
|  | Prior<br>mean /<br>Skeleton | Favored<br>dose | 0.1 | 55.38 (6.28) | 48.96 (9.07) | 23.54 (0.45) | 24.92 (0.83) | 0.34 | 0.23 | 0.18 | 0.35 | 0.33 | 0.20 |
|  |  |  | 0.25 | 71.96 (6.36) | 58.89 (9.22) | 22.99 (0.43) | 24.52 (0.88) | 0.24 | 0.18 | 0.19 | 0.36 | 0.11 | 0.06 |
|  |  | Anti-cover of<br>favored | 0.1 | 49.49 (5.49) | 48.86 (7.78) | 23.62 (0.44) | 24.77 (0.82) | 0.42 | 0.24 | 0.15 | 0.33 | 0.44 | 0.22 |
|  |  |  | 0.25 | 69.13 (5.86) | 58.56 (8.16) | 23.37 (0.40) | 24.14 (0.80) | 0.31 | 0.18 | 0.16 | 0.34 | 0.18 | 0.09 |
|  |  | Cover<br>dose of<br>favored | 0.1 | 50.91 (6.88) | 46.73 (8.86) | 23.88 (0.48) | 24.99 (0.88) | 0.31 | 0.22 | 0.25 | 0.38 | 0.31 | 0.19 |
|  |  |  | 0.25 | 66.52 (7.63) | 55.22 (8.97) | 23.43 (0.45) | 24.53 (0.91) | 0.22 | 0.17 | 0.25 | 0.40 | 0.10 | 0.06 |

## 4. Discussion

BOLD incorporates Beta DLT rate posteriors into all decision-making processes, and employs order constraints throughout each stage. Wang and Ivanova (2014) employed constraints at each stage across sampled dose-level means as well in a Bayesian continuous response model,^24^ and their approach involves simulating order-constrained posteriors using PAVA for each sampled set of values. We instead only apply PAVA to the dose-level posterior probability values for the event that the DLT rate exceeds threshold (CPATs), as in (1). Importantly, this provides a computational advantage. PAVA is only applied to a minimum set of values, and extensive simulation is averted. Thus, computations are quite fast. Note that CPAT values encompass information from the full posterior distribution in the sense that all of the posterior mass is reflected (the complement is known as well). This appears to be more informative than posterior probabilities of an interval around the target, which reflect only a portion of the posterior mass, such as in Keyboard-based methods.^11,12^ Hence, our approach is novel.

### Delineating performance differences

A series of simulations under different conditions have been studied to delineate scenarios where there are clear accuracy and efficiency differences between the proposed BOLD approach versus established methods, BOIN and CRM. There are many design parameters for Phase I trials, so we acknowledge that there are a number of parameter combinations that we have not incorporated into the scope of this work. Focus has been given on small to moderate sample size ceilings, *N_max_* ≤ 30, which we believe is a sweet spot for most investigator initiated trials. We also consider only the lowest dose as the first dose, which is the safest choice in terms of minimizing overdosing risk.

There are certain key conditions that distinguish BOLD’s performance relative to BOIN: higher accuracy at higher dose levels, more balanced accuracy across dose levels from a maximin perspective, improved relative accuracy when discrimination of the MTD versus other doses is greater (as reflected by upper *δ* values), more consistent performance as reflected by the smaller standard deviation in accuracy rate estimation, the stability of the designs in terms of smaller standard deviation of the number of patients, and the more efficient leveraging of informative prior information to improve performance, including for a simple titration design.

In informative contexts, BOLD involves specifying prior Beta probability distributions on specific DLT rate values, which is intuitive and in line with formal Bayesian analysis. In contrast, iBOIN uses skeletons with PESS values to reflect prior information, and prior probability distributions are not explicitly specified. Skeletons affect the decision boundaries for the empirical dose-level proportion. The benefits in accuracy for BOLD are apparent (Tables 2-3 and Web Tables 7-10).

A situation where BOIN clearly performs better is in identifying when all dose levels exceed MTD. By making escalation/de-escalation decisions based only on one dose level’s toxicity data, this can lead to slower dose switching, and helps in correctly identifying that dose 1 may be too toxic. When the lattice sizes are smaller, overall average accuracy is similar between BOLD and BOIN. For the select averages that exclude the bottom state, BOLD still has an advantage. In terms of the number of required patients, there is no clear advantage between BOLD and BOIN when considering all settings (Web Figure 2 and Web Tables 1-2). Any differences in mean patient number are not large, and depend on which dose level is MTD, the toxicity target, and *N_max_*.

The CRM comparison is not as extensive, and we note that a limitation of our analysis is that only the one-parameter CRM model was fit. For non-informative priors, it appears that BOLD consistently outperforms this CRM model in terms of overall average accuracy, and in a maximin sense across the dose-level accuracy rates. On the other hand, the average number of patients for CRM can be lower as the target *φ* value increases (Web Tables 1-2). Overdose rates appear comparable. Given the consistent accuracy advantage, a critical criterion, we believe that the use of BOLD is appealing.

### Overdose control

Overdosing, underestimation, and overestimation rates were monitored. In particular, as noted, the overdosing rate is higher for BOLD, especially for certain situations such as when *φ* = 0.30. Interestingly, when *τ* = 0.50 and *φ* = 0.3, if one patient in an initial cohort of 3 has a DLT at dose 1, *CPAT*_1_ = 0.49. After order constraints are applied, *PPAT*_1_ = *PPAT*_2_ = 0.49 *<* 0.50, so escalation to dose 2 is selected at the next stage. Setting *τ* = 0.48, while the PPAT values don’t change, now by the selection criterion dose 1 is maintained. The PPAT threshold parameter value can thus be adjusted to alter key dose selection decisions that impact overdosing. Note that for *φ* = 0.25, this approach has less dramatic impact, as the first dose will already be maintained after 1 out of 3 patients has a DLT after stage 1, or 1 or 2 out of 6 patients after stage 2. In terms of overall accuracy after adjustment of *τ*, note that accuracy for the higher dose levels decreases. Still, overall accuracy is only slightly affected downwards.

### Recommendations

In sum, a general set of recommendations that can be gleaned from the scenarios we considered are: 1) If there is an elevated concern that every one of the dose DLT rates could be above target (i.e. the bottom state is true), BOIN can detect this situation more accurately and quickly. If there is a belief that the MTD may reside among higher dose levels, or that at least one of the dose level DLT rates is below or equal to target rate, then BOLD is the preferable choice. 3) For smaller lattices, there is less difference in average accuracy, although this is due in part to the greater weight placed on the bottom state, when BOIN does better. 4) For smaller sample sizes, BOLD is generally more accurate, as it can switch dose levels faster. As the maximum sample size is set higher, the accuracy gap can decrease somewhat, but adjustments to 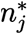 values can help maintain BOLD’s accuracy advantage. 5) If the MTD is clearly distinguished in terms of the difference in DLT rate values from the dose above it, BOLD can leverage this more effectively to improve accuracy. If the MTD is not well distinguished, all the methods considered here do not perform well. 6) BOIN has less overdosing. However, this is in part due to a trade-off of BOLD having higher accuracy at higher dose levels. Overdose control through adjusting the PPAT threshold parameter *τ* can be helpful, depending on *φ* and the prior specifications for DLT rate parameters. 7) If it is of interest to incorporate prior information, BOLD can perform decisively better.

We encourage the use of the respective R codes to help guide design adoption and inputs. In a Bayes risk sense, averages of dose-level accuracy and other performance statistics can be weighted by prior belief as to which dose level is the MTD. Note that our selective averaging that excludes the bottom state is an example of this approach.

## 6. Conclusion

We propose a new and computationally fast approach that relies on prior distribution specifications at the DLT rate parameter level. The primary characteristic of the proposed Bayesian Ordered Lattice Design (BOLD) is its integration of posterior information through order constraints that are imposed at each decision stage. This enables “shared learning” about toxicity across dose levels. We delineate when advantages through improved accuracy in MTD identification can arise in Bayesian dose finding trials.

The posterior probability of fundamental interest relates to whether a dose DLT rate exceeds the toxicity threshold. BOLD focuses on this posterior probability as a criterion for both dose selection and early stopping. Extensive simulation studies support that the accuracy performance of BOLD can often surpass BOIN and CRM. In an adaptation for titration and when Bayesian informative priors are utilized, BOLD demonstrates superior performance. Lastly, the BOLD framework is flexible, so the proposed methods can be extended to more general Bayesian phase I design settings that incorporate more complex information. Such designs will be elaborated upon in forthcoming work.

## Data Availability

All data produced in the present work are contained in the manuscript

## Acknowledgements

This work was supported in part by grants NSF 2333326, P30 CA134274-10 (University of Maryland, Baltimore), and P30 CA043703-34 (Case Comprehensive Cancer Center).

## Supplementary Material

**Web Figure 1:**
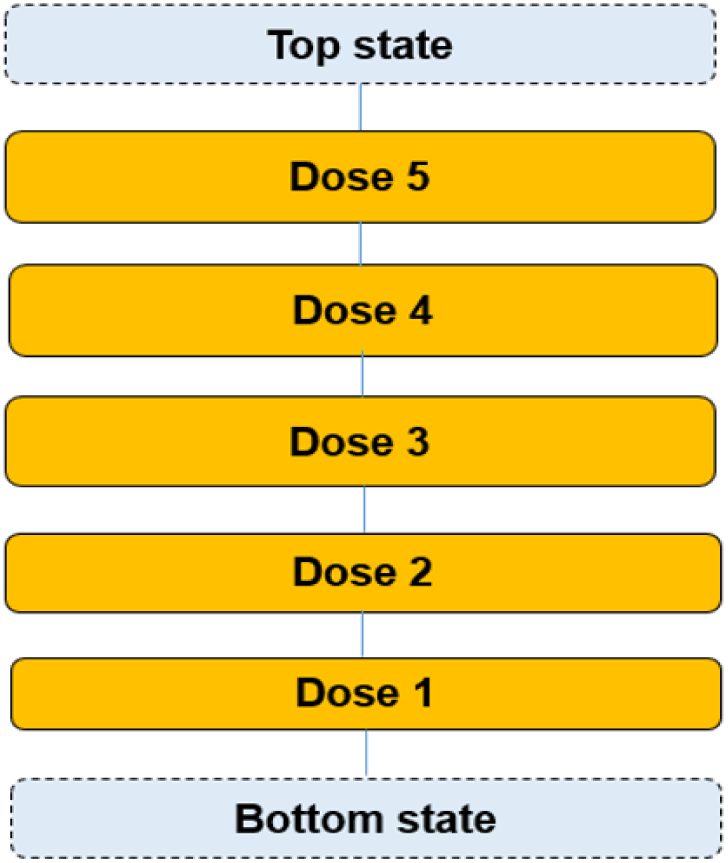
Hasse diagram of 5-dose lattice with linear order, where the bottom state represents that a MTD is below the minimum dose, and the top state signifies that the MTD exceeds the maximum dose. Classification is to one state, and classification to a specific dose level indicates that the corresponding dose is identified as the MTD.

**Web Figure 2:**
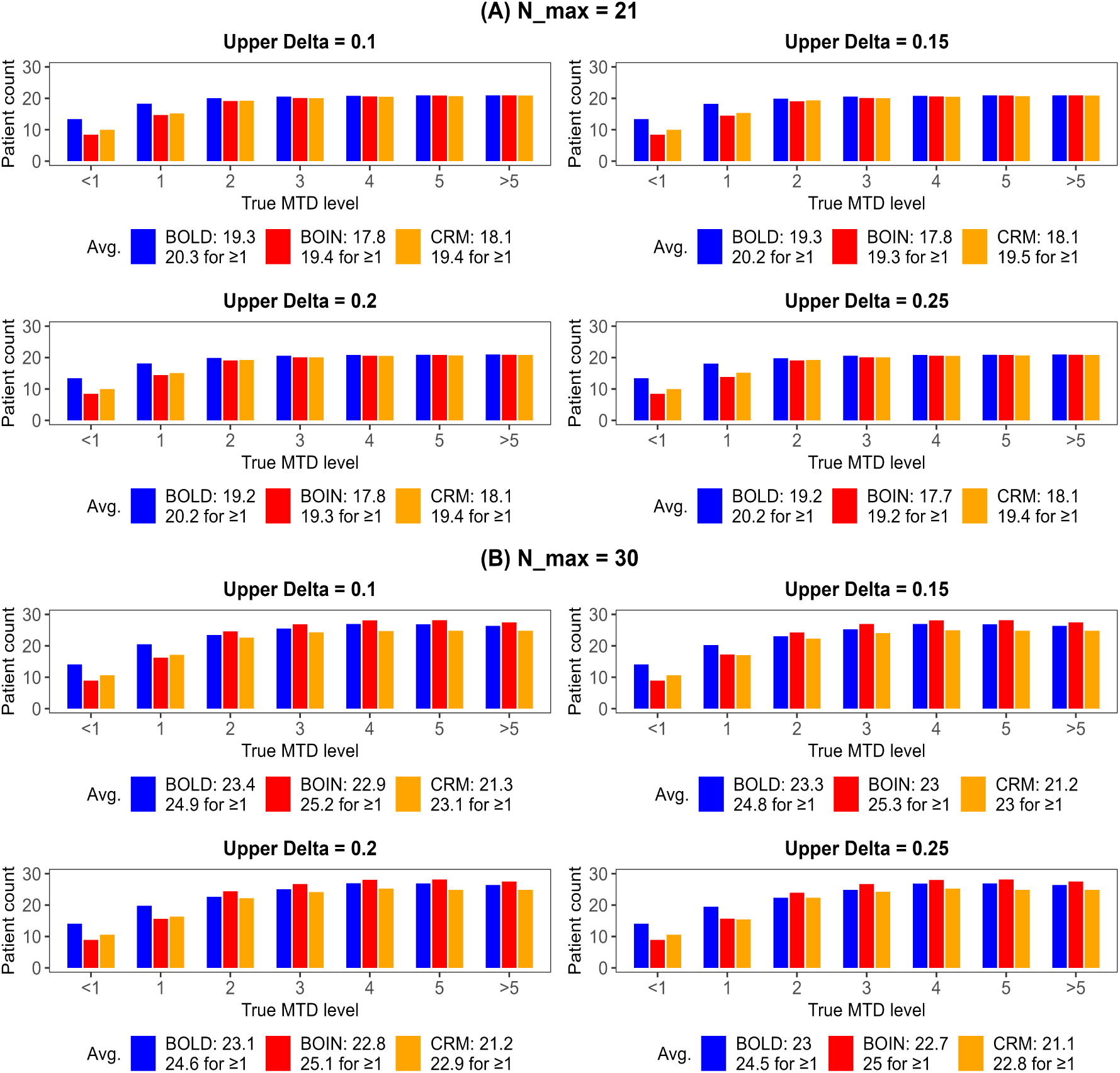
Efficiency of BOLD, BOIN and CRM design, represented by the average patient count, at 5-dose model with non-informative prior by MTD level and stratified by *N_max_* and upper *δ*, assuming random scenarios of true DLT rates, at target rate *φ* = 0.25. The legend denotes the average for all the 7 MTD levels (1st row) and for the MTD levels that are *≥* 1 (2nd row).

**Web Figure 3:**
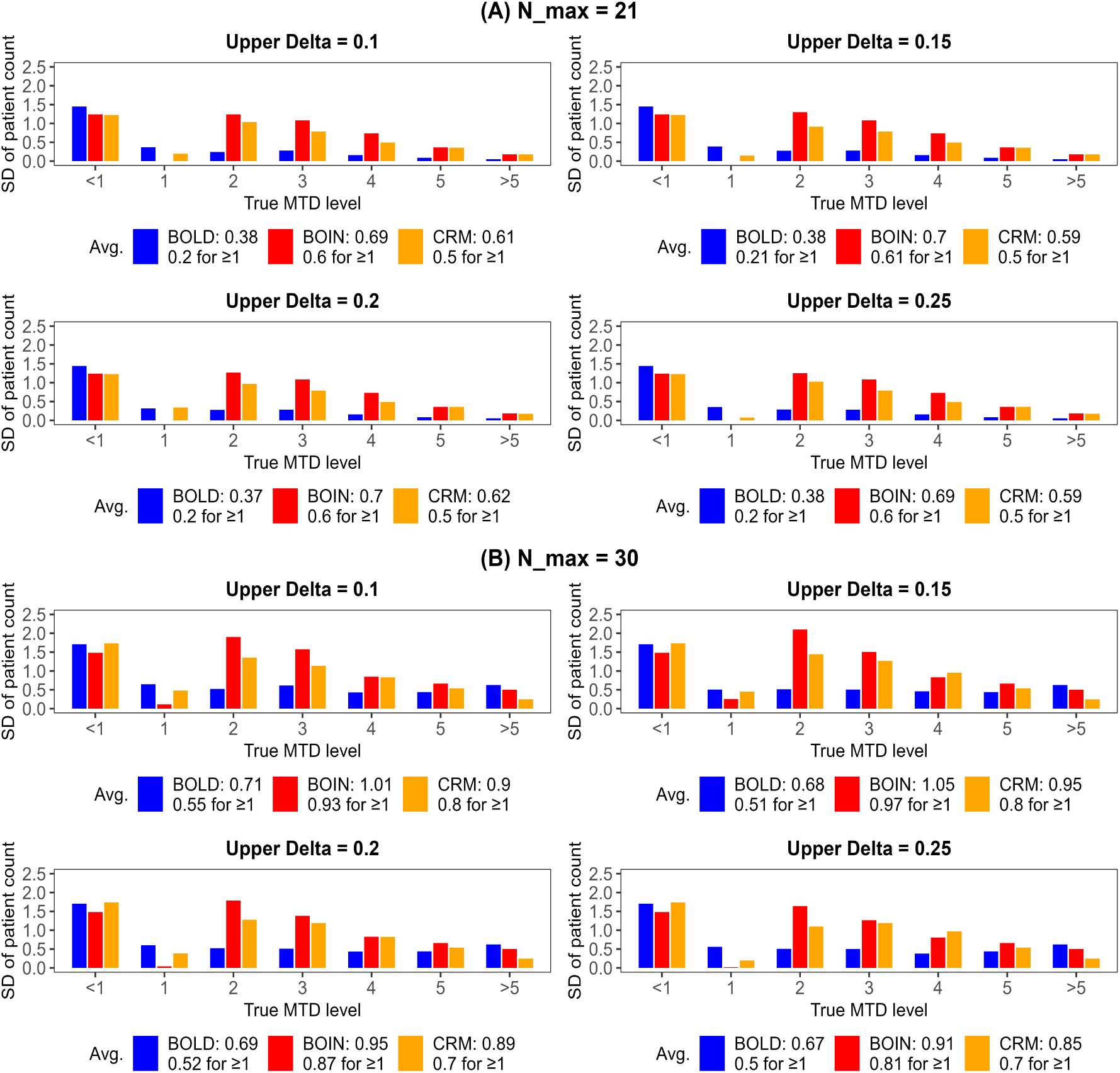
Standard deviation (SD) of patient count of BOLD, BOIN and CRM design across scenarios at 5-dose model with non-informative prior by MTD level and stratified by *N_max_* and upper *δ*, assuming random scenarios of true DLT rates, at target rate *φ* = 0.25. The legend denotes the average for all the 7 MTD levels (1st row) and for the MTD levels that are *≥* 1 (2nd row).

**Web Figure 4:**
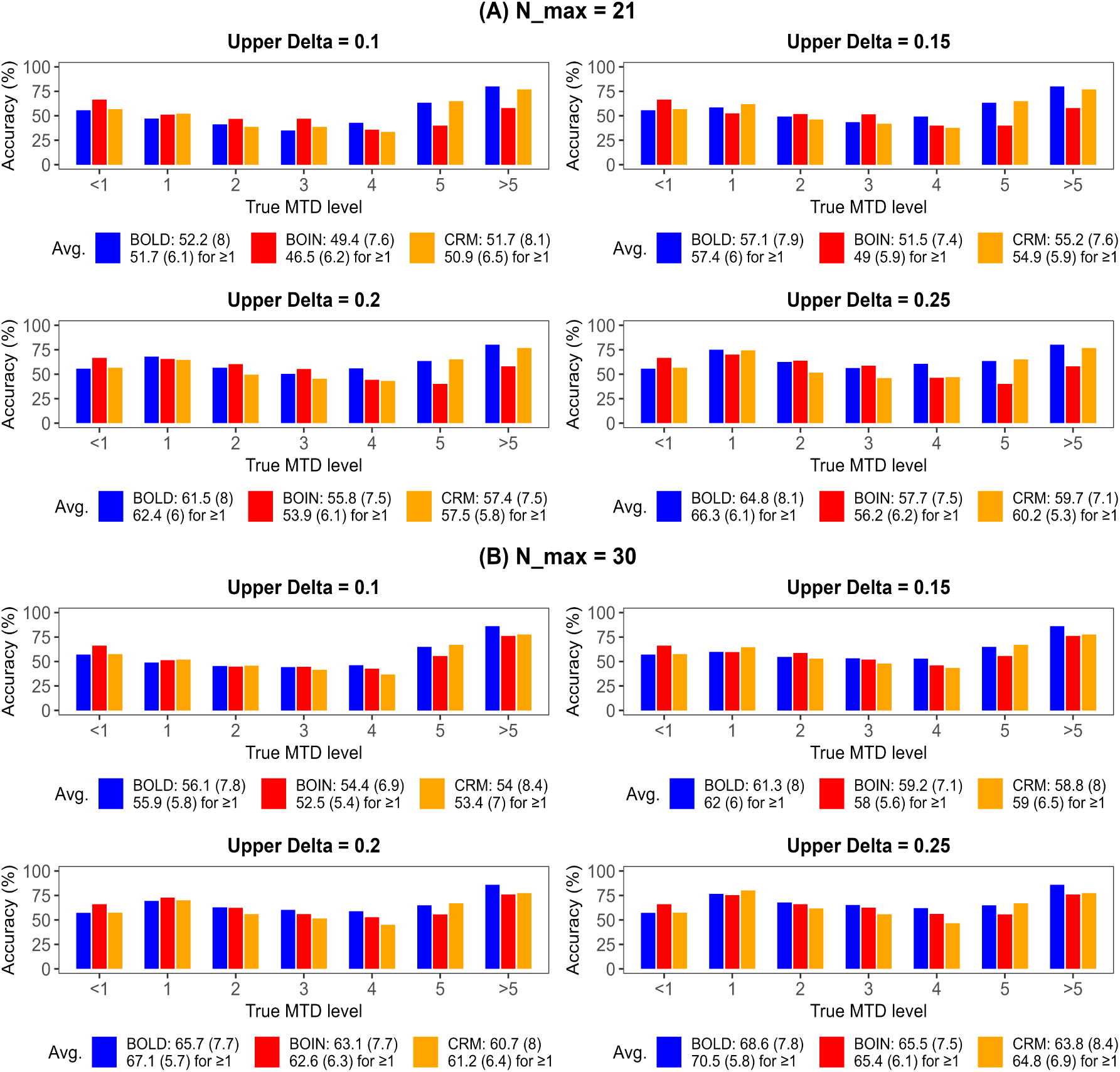
Accuracy of BOLD, BOIN, and CRM design, represented by the average of accurate MTD identifications (%) across scenarios, at 5-dose model with non-informative prior by MTD level and stratified by *N_max_* and upper *δ*, assuming random scenarios of true DLT rates, at target rate *φ* = 0.15. The legend denotes the average and SD (in parenthesis) for all the 7 MTD levels (1st row) and for the MTD levels that are *≥* 1 (2nd row).

**Web Figure 5:**
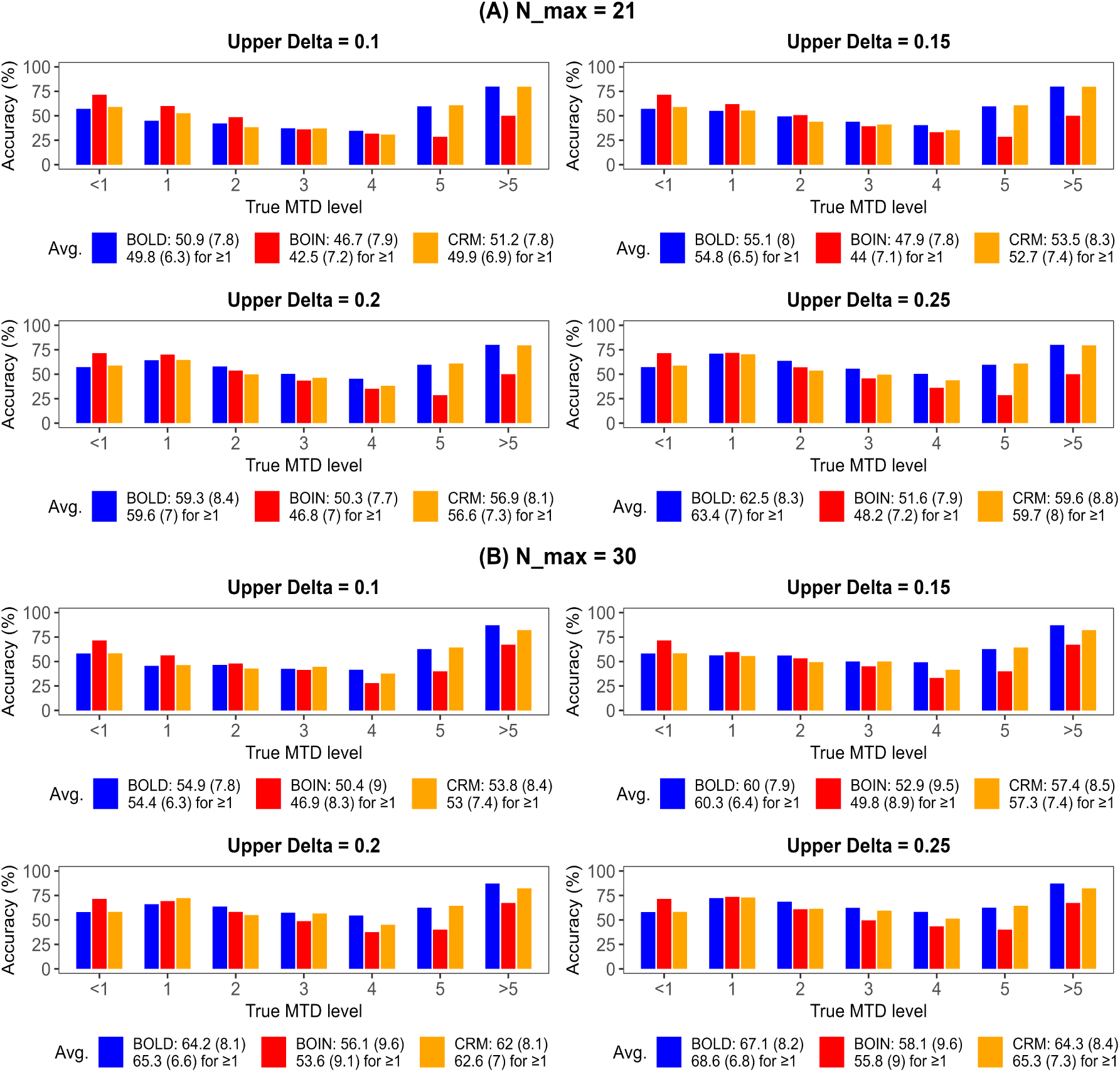
Accuracy of BOLD, BOIN, and CRM design, represented by the average of accurate MTD identifications (%) across scenarios, at 5-dose model with non-informative prior by MTD level and stratified by *N_max_* and upper *δ*, assuming random scenarios of true DLT rates, at target rate *φ* = 0.2. The legend denotes the average and SD (in parenthesis) for all the 7 MTD levels (1st row) and for the MTD levels that are *≥* 1 (2nd row).

**Web Figure 6:**
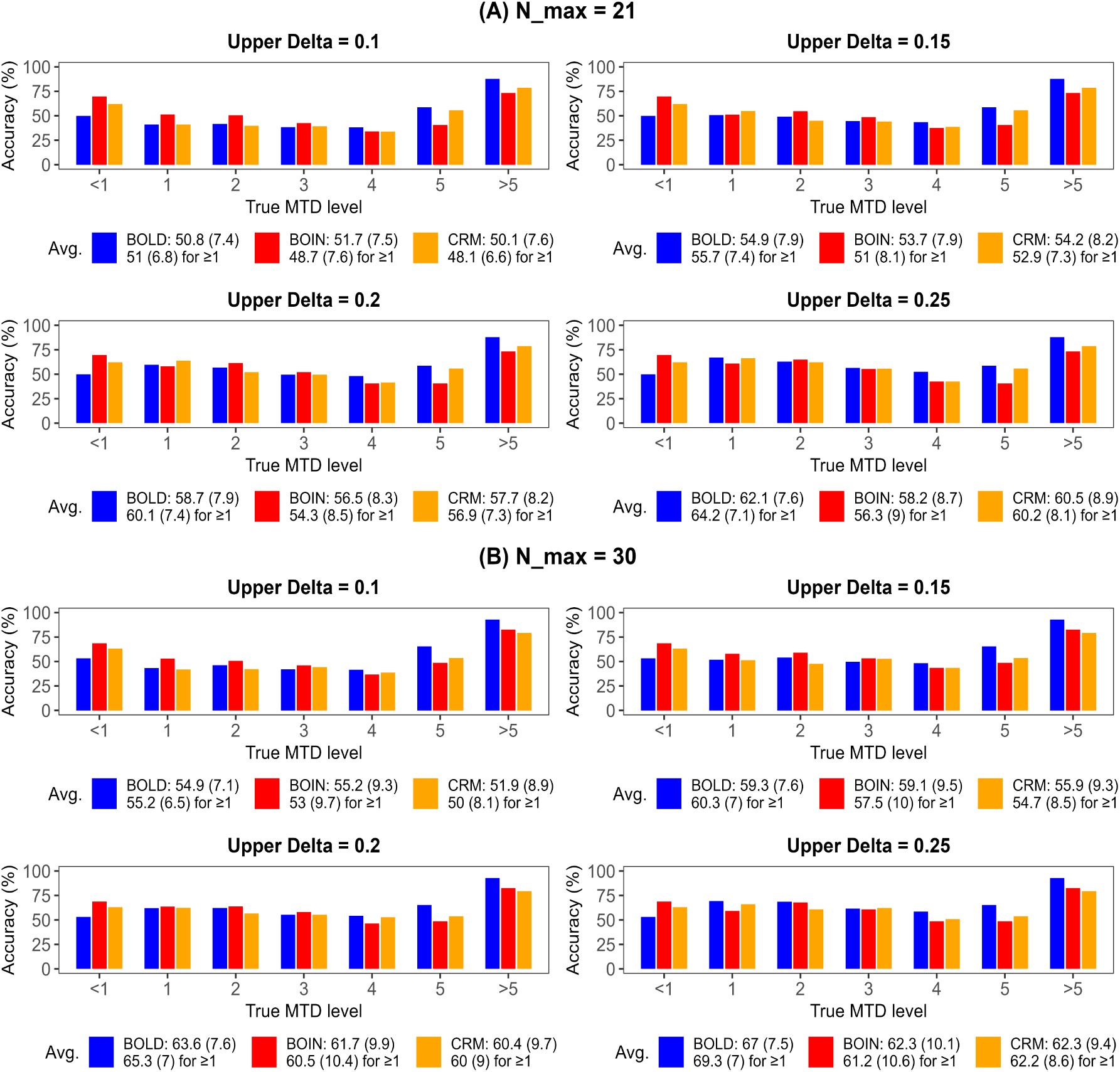
Accuracy of BOLD, BOIN, and CRM design, represented by the average of accurate MTD identifications (%) across scenarios, at 5-dose model with non-informative prior by MTD level and stratified by *N_max_* and upper *δ*, assuming random scenarios of true DLT rates, at target rate *φ* = 0.3. The legend denotes the average and SD (in parenthesis) for all the 7 MTD levels (1st row) and for the MTD levels that are *≥* 1 (2nd row).

**Web Table 1:**
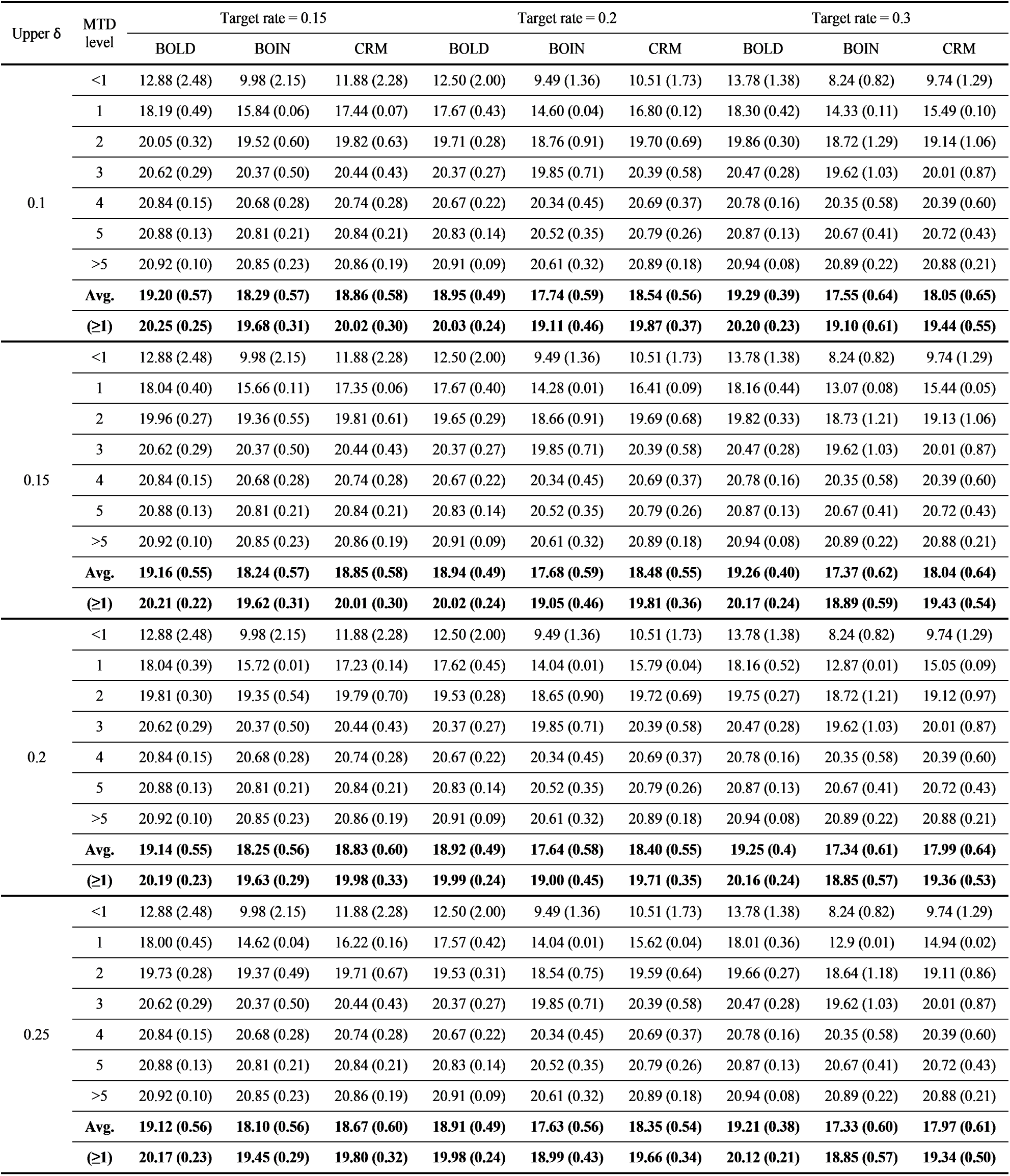
Summary of efficiency, represented by the average patient count and its standard deviation (in parenthesis), of BOLD, BOIN and CRM by upper *δ* value of random scenarios for *φ* = 0.15, 0.2 and 0.3 and *N_max_* = 21 at 5-dose model with non-informative prior, where “Avg.” denotes the average for all the 7 MTD levels and “(*≥*1)” denotes the average for the MTD levels that are *≥* 1.

| Upper $\delta$ | MTD level | Target rate = 0.15 | | | Target rate = 0.2 | | | Target rate = 0.3 | | |
| --- | --- | --- | --- | --- | --- | --- | --- | --- | --- | --- |
|  |  | BOLD | BOIN | CRM | BOLD | BOIN | CRM | BOLD | BOIN | CRM |
| 0.1 | <1 | 12.88 (2.48) | 9.98 (2.15) | 11.88 (2.28) | 12.50 (2.00) | 9.49 (1.36) | 10.51 (1.73) | 13.78 (1.38) | 8.24 (0.82) | 9.74 (1.29) |
|  | 1 | 18.19 (0.49) | 15.84 (0.06) | 17.44 (0.07) | 17.67 (0.43) | 14.60 (0.04) | 16.80 (0.12) | 18.30 (0.42) | 14.33 (0.11) | 15.49 (0.10) |
|  | 2 | 20.05 (0.32) | 19.52 (0.60) | 19.82 (0.63) | 19.71 (0.28) | 18.76 (0.91) | 19.70 (0.69) | 19.86 (0.30) | 18.72 (1.29) | 19.14 (1.06) |
|  | 3 | 20.62 (0.29) | 20.37 (0.50) | 20.44 (0.43) | 20.37 (0.27) | 19.85 (0.71) | 20.39 (0.58) | 20.47 (0.28) | 19.62 (1.03) | 20.01 (0.87) |
|  | 4 | 20.84 (0.15) | 20.68 (0.28) | 20.74 (0.28) | 20.67 (0.22) | 20.34 (0.45) | 20.69 (0.37) | 20.78 (0.16) | 20.35 (0.58) | 20.39 (0.60) |
|  | 5 | 20.88 (0.13) | 20.81 (0.21) | 20.84 (0.21) | 20.83 (0.14) | 20.52 (0.35) | 20.79 (0.26) | 20.87 (0.13) | 20.67 (0.41) | 20.72 (0.43) |
|  | >5 | 20.92 (0.10) | 20.85 (0.23) | 20.86 (0.19) | 20.91 (0.09) | 20.61 (0.32) | 20.89 (0.18) | 20.94 (0.08) | 20.89 (0.22) | 20.88 (0.21) |
|  | **Avg.** | **19.20 (0.57)** | **18.29 (0.57)** | **18.86 (0.58)** | **18.95 (0.49)** | **17.74 (0.59)** | **18.54 (0.56)** | **19.29 (0.39)** | **17.55 (0.64)** | **18.05 (0.65)** |
|  | **($\geq 1$)** | **20.25 (0.25)** | **19.68 (0.31)** | **20.02 (0.30)** | **20.03 (0.24)** | **19.11 (0.46)** | **19.87 (0.37)** | **20.20 (0.23)** | **19.10 (0.61)** | **19.44 (0.55)** |
| 0.15 | <1 | 12.88 (2.48) | 9.98 (2.15) | 11.88 (2.28) | 12.50 (2.00) | 9.49 (1.36) | 10.51 (1.73) | 13.78 (1.38) | 8.24 (0.82) | 9.74 (1.29) |
|  | 1 | 18.04 (0.40) | 15.66 (0.11) | 17.35 (0.06) | 17.67 (0.40) | 14.28 (0.01) | 16.41 (0.09) | 18.16 (0.44) | 13.07 (0.08) | 15.44 (0.05) |
|  | 2 | 19.96 (0.27) | 19.36 (0.55) | 19.81 (0.61) | 19.65 (0.29) | 18.66 (0.91) | 19.69 (0.68) | 19.82 (0.33) | 18.73 (1.21) | 19.13 (1.06) |
|  | 3 | 20.62 (0.29) | 20.37 (0.50) | 20.44 (0.43) | 20.37 (0.27) | 19.85 (0.71) | 20.39 (0.58) | 20.47 (0.28) | 19.62 (1.03) | 20.01 (0.87) |
|  | 4 | 20.84 (0.15) | 20.68 (0.28) | 20.74 (0.28) | 20.67 (0.22) | 20.34 (0.45) | 20.69 (0.37) | 20.78 (0.16) | 20.35 (0.58) | 20.39 (0.60) |
|  | 5 | 20.88 (0.13) | 20.81 (0.21) | 20.84 (0.21) | 20.83 (0.14) | 20.52 (0.35) | 20.79 (0.26) | 20.87 (0.13) | 20.67 (0.41) | 20.72 (0.43) |
|  | >5 | 20.92 (0.10) | 20.85 (0.23) | 20.86 (0.19) | 20.91 (0.09) | 20.61 (0.32) | 20.89 (0.18) | 20.94 (0.08) | 20.89 (0.22) | 20.88 (0.21) |
|  | **Avg.** | **19.16 (0.55)** | **18.24 (0.57)** | **18.85 (0.58)** | **18.94 (0.49)** | **17.68 (0.59)** | **18.48 (0.55)** | **19.26 (0.40)** | **17.37 (0.62)** | **18.04 (0.64)** |
|  | **($\geq 1$)** | **20.21 (0.22)** | **19.62 (0.31)** | **20.01 (0.30)** | **20.02 (0.24)** | **19.05 (0.46)** | **19.81 (0.36)** | **20.17 (0.24)** | **18.89 (0.59)** | **19.43 (0.54)** |
| 0.2 | <1 | 12.88 (2.48) | 9.98 (2.15) | 11.88 (2.28) | 12.50 (2.00) | 9.49 (1.36) | 10.51 (1.73) | 13.78 (1.38) | 8.24 (0.82) | 9.74 (1.29) |
|  | 1 | 18.04 (0.39) | 15.72 (0.01) | 17.23 (0.14) | 17.62 (0.45) | 14.04 (0.01) | 15.79 (0.04) | 18.16 (0.52) | 12.87 (0.01) | 15.05 (0.09) |
|  | 2 | 19.81 (0.30) | 19.35 (0.54) | 19.79 (0.70) | 19.53 (0.28) | 18.65 (0.90) | 19.72 (0.69) | 19.75 (0.27) | 18.72 (1.21) | 19.12 (0.97) |
|  | 3 | 20.62 (0.29) | 20.37 (0.50) | 20.44 (0.43) | 20.37 (0.27) | 19.85 (0.71) | 20.39 (0.58) | 20.47 (0.28) | 19.62 (1.03) | 20.01 (0.87) |
|  | 4 | 20.84 (0.15) | 20.68 (0.28) | 20.74 (0.28) | 20.67 (0.22) | 20.34 (0.45) | 20.69 (0.37) | 20.78 (0.16) | 20.35 (0.58) | 20.39 (0.60) |
|  | 5 | 20.88 (0.13) | 20.81 (0.21) | 20.84 (0.21) | 20.83 (0.14) | 20.52 (0.35) | 20.79 (0.26) | 20.87 (0.13) | 20.67 (0.41) | 20.72 (0.43) |
|  | >5 | 20.92 (0.10) | 20.85 (0.23) | 20.86 (0.19) | 20.91 (0.09) | 20.61 (0.32) | 20.89 (0.18) | 20.94 (0.08) | 20.89 (0.22) | 20.88 (0.21) |
|  | **Avg.** | **19.14 (0.55)** | **18.25 (0.56)** | **18.83 (0.60)** | **18.92 (0.49)** | **17.64 (0.58)** | **18.40 (0.55)** | **19.25 (0.4)** | **17.34 (0.61)** | **17.99 (0.64)** |
|  | **($\geq 1$)** | **20.19 (0.23)** | **19.63 (0.29)** | **19.98 (0.33)** | **19.99 (0.24)** | **19.00 (0.45)** | **19.71 (0.35)** | **20.16 (0.24)** | **18.85 (0.57)** | **19.36 (0.53)** |
| 0.25 | <1 | 12.88 (2.48) | 9.98 (2.15) | 11.88 (2.28) | 12.50 (2.00) | 9.49 (1.36) | 10.51 (1.73) | 13.78 (1.38) | 8.24 (0.82) | 9.74 (1.29) |
|  | 1 | 18.00 (0.45) | 14.62 (0.04) | 16.22 (0.16) | 17.57 (0.42) | 14.04 (0.01) | 15.62 (0.04) | 18.01 (0.36) | 12.9 (0.01) | 14.94 (0.02) |
|  | 2 | 19.73 (0.28) | 19.37 (0.49) | 19.71 (0.67) | 19.53 (0.31) | 18.54 (0.75) | 19.59 (0.64) | 19.66 (0.27) | 18.64 (1.18) | 19.11 (0.86) |
|  | 3 | 20.62 (0.29) | 20.37 (0.50) | 20.44 (0.43) | 20.37 (0.27) | 19.85 (0.71) | 20.39 (0.58) | 20.47 (0.28) | 19.62 (1.03) | 20.01 (0.87) |
|  | 4 | 20.84 (0.15) | 20.68 (0.28) | 20.74 (0.28) | 20.67 (0.22) | 20.34 (0.45) | 20.69 (0.37) | 20.78 (0.16) | 20.35 (0.58) | 20.39 (0.60) |
|  | 5 | 20.88 (0.13) | 20.81 (0.21) | 20.84 (0.21) | 20.83 (0.14) | 20.52 (0.35) | 20.79 (0.26) | 20.87 (0.13) | 20.67 (0.41) | 20.72 (0.43) |
|  | >5 | 20.92 (0.10) | 20.85 (0.23) | 20.86 (0.19) | 20.91 (0.09) | 20.61 (0.32) | 20.89 (0.18) | 20.94 (0.08) | 20.89 (0.22) | 20.88 (0.21) |
|  | **Avg.** | **19.12 (0.56)** | **18.10 (0.56)** | **18.67 (0.60)** | **18.91 (0.49)** | **17.63 (0.56)** | **18.35 (0.54)** | **19.21 (0.38)** | **17.33 (0.60)** | **17.97 (0.61)** |
|  | **($\geq 1$)** | **20.17 (0.23)** | **19.45 (0.29)** | **19.80 (0.32)** | **19.98 (0.24)** | **18.99 (0.43)** | **19.66 (0.34)** | **20.12 (0.21)** | **18.85 (0.57)** | **19.34 (0.50)** |

**Web Table 2:** Summary of efficiency, represented by the average patient count and its standard deviation (in parenthesis), of BOLD, BOIN and CRM by upper *δ* value of random scenarios for *φ* = 0.15, 0.2 and 0.3 and *N_max_* = 30 at 5-dose model with non-informative prior, where “Avg.” denotes the average for all the 7 MTD levels and “(*≥*1)” denotes the average for the MTD levels that are *≥* 1.

| Upper $\delta$ | MTD level | Target rate = 0.15 | | | Target rate = 0.2 | | | Target rate = 0.3 | | |
| --- | --- | --- | --- | --- | --- | --- | --- | --- | --- | --- |
|  |  | BOLD | BOIN | CRM | BOLD | BOIN | CRM | BOLD | BOIN | CRM |
| 0.1 | <1 | 13.76 (2.84) | 10.52 (2.45) | 12.68 (2.68) | 13.12 (2.32) | 9.83 (1.58) | 11.46 (2.19) | 14.98 (1.66) | 8.70 (0.87) | 9.93 (1.45) |
|  | 1 | 20.43 (0.62) | 17.21 (0.21) | 19.44 (0.44) | 19.55 (0.55) | 15.86 (0.12) | 18.39 (0.39) | 21.13 (0.58) | 15.50 (0.17) | 16.19 (0.35) |
|  | 2 | 23.40 (0.52) | 23.52 (0.71) | 24.26 (0.79) | 22.71 (0.49) | 22.29 (1.74) | 23.27 (0.83) | 23.69 (0.58) | 21.52 (1.58) | 21.80 (1.18) |
|  | 3 | 25.35 (0.50) | 25.40 (0.82) | 25.95 (0.78) | 24.61 (0.46) | 24.43 (1.33) | 25.22 (0.99) | 25.00 (0.66) | 24.15 (1.30) | 23.48 (1.38) |
|  | 4 | 26.13 (0.49) | 26.83 (0.67) | 26.42 (0.78) | 25.70 (0.55) | 25.59 (0.82) | 25.80 (0.64) | 25.76 (0.45) | 25.38 (1.05) | 24.45 (0.81) |
|  | 5 | 25.80 (0.30) | 26.86 (0.37) | 25.73 (0.33) | 25.85 (0.37) | 26.17 (0.78) | 25.51 (0.42) | 25.51 (0.43) | 26.45 (0.91) | 24.87 (0.73) |
|  | >5 | 25.39 (0.36) | 26.18 (0.35) | 25.06 (0.35) | 25.66 (0.42) | 25.86 (0.51) | 25.00 (0.31) | 24.63 (0.32) | 26.01 (0.47) | 24.90 (0.32) |
|  | Avg. | **22.89 (0.81)** | **22.36 (0.80)** | **22.79 (0.88)** | **22.46 (0.74)** | **21.43 (0.98)** | **22.09 (0.82)** | **22.96 (0.67)** | **21.10 (0.91)** | **20.80 (0.89)** |
| | ($\geq 1$) | **24.42 (0.47)** | **24.33 (0.52)** | **24.48 (0.58)** | **24.01 (0.47)** | **23.37 (0.88)** | **23.86 (0.60)** | **24.29 (0.50)** | **23.17 (0.92)** | **22.62 (0.79)** |
| 0.15 | <1 | 13.76 (2.84) | 10.52 (2.45) | 12.68 (2.68) | 13.12 (2.32) | 9.83 (1.58) | 11.46 (2.19) | 14.98 (1.66) | 8.70 (0.87) | 9.93 (1.45) |
|  | 1 | 20.02 (0.66) | 16.46 (0.08) | 18.51 (0.18) | 19.31 (0.65) | 15.87 (0.19) | 18.27 (0.31) | 20.77 (0.71) | 14.32 (0.11) | 16.73 (0.32) |
|  | 2 | 23.07 (0.55) | 22.98 (0.60) | 24.12 (1.06) | 22.38 (0.57) | 21.63 (1.66) | 23.24 (1.02) | 23.49 (0.56) | 21.27 (1.51) | 21.34 (1.41) |
|  | 3 | 25.20 (0.53) | 25.39 (0.86) | 25.97 (0.89) | 24.27 (0.48) | 24.00 (1.24) | 25.23 (1.02) | 24.94 (0.70) | 23.87 (1.29) | 23.30 (1.18) |
|  | 4 | 26.39 (0.43) | 27.06 (0.66) | 26.67 (0.71) | 25.79 (0.55) | 25.79 (0.93) | 25.93 (0.58) | 26.02 (0.50) | 25.57 (1.13) | 24.43 (0.74) |
|  | 5 | 25.80 (0.30) | 26.86 (0.37) | 25.73 (0.33) | 25.85 (0.37) | 26.17 (0.78) | 25.51 (0.42) | 25.51 (0.43) | 26.45 (0.91) | 24.87 (0.73) |
|  | >5 | 25.39 (0.36) | 26.18 (0.35) | 25.06 (0.35) | 25.66 (0.42) | 25.86 (0.51) | 25.00 (0.31) | 24.63 (0.32) | 26.01 (0.47) | 24.90 (0.32) |
|  | Avg. | **22.80 (0.81)** | **22.20 (0.77)** | **22.68 (0.88)** | **22.34 (0.77)** | **21.31 (0.98)** | **22.09 (0.84)** | **22.9 (0.70)** | **20.88 (0.90)** | **20.79 (0.88)** |
| | ($\geq 1$) | **24.31 (0.47)** | **24.15 (0.49)** | **24.35 (0.58)** | **23.88 (0.51)** | **23.22 (0.89)** | **23.86 (0.61)** | **24.23 (0.54)** | **22.92 (0.90)** | **22.60 (0.78)** |
| 0.2 | <1 | 13.76 (2.84) | 10.52 (2.45) | 12.68 (2.68) | 13.12 (2.32) | 9.83 (1.58) | 11.46 (2.19) | 14.98 (1.66) | 8.70 (0.87) | 9.93 (1.45) |
|  | 1 | 19.79 (0.60) | 16.19 (0.12) | 18.57 (0.47) | 19.36 (0.68) | 14.61 (0.01) | 18.50 (0.37) | 20.53 (0.77) | 14.58 (0.01) | 16.92 (0.25) |
|  | 2 | 22.73 (0.54) | 22.31 (0.54) | 23.67 (1.07) | 22.03 (0.47) | 21.41 (1.45) | 23.10 (1.03) | 23.19 (0.56) | 21.42 (1.43) | 21.31 (1.30) |
|  | 3 | 25.01 (0.43) | 25.09 (0.82) | 26.18 (0.92) | 24.23 (0.44) | 23.93 (1.48) | 25.47 (0.89) | 24.95 (0.66) | 23.66 (1.20) | 23.24 (1.30) |
|  | 4 | 26.54 (0.43) | 27.21 (0.52) | 26.81 (0.71) | 25.93 (0.53) | 25.91 (0.83) | 26.00 (0.53) | 26.14 (0.48) | 25.53 (1.22) | 24.77 (0.79) |
|  | 5 | 25.80 (0.30) | 26.86 (0.37) | 25.73 (0.33) | 25.85 (0.37) | 26.17 (0.78) | 25.51 (0.42) | 25.51 (0.43) | 26.45 (0.91) | 24.87 (0.73) |
|  | >5 | 25.39 (0.36) | 26.18 (0.35) | 25.06 (0.35) | 25.66 (0.42) | 25.86 (0.51) | 25.00 (0.31) | 24.63 (0.32) | 26.01 (0.47) | 24.90 (0.32) |
|  | Avg. | **22.72 (0.79)** | **22.05 (0.74)** | **22.67 (0.93)** | **22.31 (0.75)** | **21.10 (0.95)** | **22.15 (0.82)** | **22.85 (0.69)** | **20.91 (0.87)** | **20.85 (0.88)** |
| | ($\geq 1$) | **24.21 (0.45)** | **23.97 (0.45)** | **24.34 (0.64)** | **23.84 (0.49)** | **22.98 (0.84)** | **23.93 (0.59)** | **24.16 (0.53)** | **22.94 (0.87)** | **22.67 (0.78)** |
| 0.25 | <1 | 13.76 (2.84) | 10.52 (2.45) | 12.68 (2.68) | 13.12 (2.32) | 9.83 (1.58) | 11.46 (2.19) | 14.98 (1.66) | 8.70 (0.87) | 9.93 (1.45) |
|  | 1 | 19.71 (0.54) | 15.48 (0.01) | 18.38 (0.53) | 19.07 (0.58) | 14.43 (0.01) | 16.79 (0.19) | 20.26 (0.66) | 13.32 (0.01) | 14.64 (0.12) |
|  | 2 | 22.32 (0.48) | 22.59 (0.54) | 23.09 (0.89) | 21.72 (0.55) | 21.21 (1.41) | 22.75 (0.89) | 22.99 (0.57) | 21.38 (1.54) | 21.19 (1.37) |
|  | 3 | 24.89 (0.38) | 25.05 (0.79) | 26.03 (0.85) | 23.94 (0.46) | 23.78 (1.49) | 25.31 (0.98) | 24.81 (0.59) | 23.50 (1.21) | 23.61 (1.24) |
|  | 4 | 26.61 (0.40) | 26.75 (0.61) | 26.78 (0.70) | 25.93 (0.46) | 25.86 (0.85) | 26.12 (0.60) | 26.16 (0.49) | 25.38 (1.22) | 24.78 (0.79) |
|  | 5 | 25.80 (0.30) | 26.86 (0.37) | 25.73 (0.33) | 25.85 (0.37) | 26.17 (0.78) | 25.51 (0.42) | 25.51 (0.43) | 26.45 (0.91) | 24.87 (0.73) |
|  | >5 | 25.39 (0.36) | 26.18 (0.35) | 25.06 (0.35) | 25.66 (0.42) | 25.86 (0.51) | 25.00 (0.31) | 24.63 (0.32) | 26.01 (0.47) | 24.90 (0.32) |
|  | Avg. | **22.64 (0.76)** | **21.92 (0.73)** | **22.54 (0.90)** | **22.18 (0.74)** | **21.02 (0.95)** | **21.85 (0.80)** | **22.76 (0.67)** | **20.68 (0.89)** | **20.56 (0.86)** |
| | ($\geq 1$) | **24.12 (0.41)** | **23.82 (0.44)** | **24.18 (0.61)** | **23.70 (0.47)** | **22.89 (0.84)** | **23.58 (0.57)** | **24.06 (0.51)** | **22.67 (0.89)** | **22.33 (0.76)** |

**Web Table 3:** Summary of overdose, underestimation, and overestimation rates of BOLD, BOIN and CRM at 5-dose model with non-informative prior for *φ* = 0.25 and 0.3 with *N_max_* = 21 and 30, assuming random scenarios of true DLT rates with upper *δ* = 0.15, where “Avg.” denotes the average for all the MTD levels and “(*≥*1)” denotes the average for the MTD levels that are *≥* 1.

| Target rate | N max | MTD level | Overdose rate |  |  | Underestimation rate |  |  | Overestimation rate |  |  |
| --- | --- | --- | --- | --- | --- | --- | --- | --- | --- | --- | --- |
|  |  |  | BOLD | BOIN | CRM | BOLD | BOIN | CRM | BOLD | BOIN | CRM |
| 0.25 | 21 | <1 | 0.21 | 0.18 | 0.14 | -- | -- | -- | 0.49 | 0.28 | 0.42 |
|  |  | 1 | 0.43 | 0.35 | 0.36 | 0.10 | 0.29 | 0.20 | 0.31 | 0.14 | 0.30 |
|  |  | 2 | 0.30 | 0.17 | 0.36 | 0.19 | 0.31 | 0.17 | 0.29 | 0.16 | 0.39 |
|  |  | 3 | 0.20 | 0.10 | 0.26 | 0.27 | 0.37 | 0.24 | 0.26 | 0.11 | 0.35 |
|  |  | 4 | 0.11 | 0.05 | 0.17 | 0.36 | 0.59 | 0.32 | 0.20 | 0.07 | 0.33 |
|  |  | 5 | -- | -- | -- | 0.50 | 0.67 | 0.39 | -- | -- | -- |
|  |  | >5 | -- | -- | -- | 0.22 | 0.35 | 0.17 | -- | -- | -- |
|  |  | Avg. | **0.25** | **0.17** | **0.26** | **0.27** | **0.43** | **0.25** | **0.31** | **0.15** | **0.36** |
| | | ($\geq 1$) | **0.26** | **0.17** | **0.29** | **0.27** | **0.43** | **0.25** | **0.26** | **0.12** | **0.34** |
|  | 30 | <1 | 0.25 | 0.19 | 0.14 | -- | -- | -- | 0.48 | 0.28 | 0.42 |
|  |  | 1 | 0.42 | 0.38 | 0.36 | 0.10 | 0.24 | 0.20 | 0.30 | 0.13 | 0.28 |
|  |  | 2 | 0.31 | 0.21 | 0.36 | 0.17 | 0.25 | 0.19 | 0.27 | 0.16 | 0.32 |
|  |  | 3 | 0.26 | 0.15 | 0.27 | 0.25 | 0.35 | 0.25 | 0.24 | 0.16 | 0.28 |
|  |  | 4 | 0.18 | 0.09 | 0.18 | 0.30 | 0.46 | 0.31 | 0.22 | 0.15 | 0.25 |
|  |  | 5 | -- | -- | -- | 0.37 | 0.53 | 0.43 | -- | -- | -- |
|  |  | >5 | -- | -- | -- | 0.09 | 0.23 | 0.19 | -- | -- | -- |
|  |  | Avg. | **0.29** | **0.20** | **0.26** | **0.21** | **0.34** | **0.26** | **0.30** | **0.17** | **0.31** |
| | | ($\geq 1$) | **0.29** | **0.21** | **0.29** | **0.21** | **0.34** | **0.26** | **0.26** | **0.15** | **0.28** |
| 0.3 | 21 | <1 | 0.36 | 0.16 | 0.11 | -- | -- | -- | 0.50 | 0.30 | 0.38 |
|  |  | 1 | 0.51 | 0.37 | 0.32 | 0.12 | 0.28 | 0.19 | 0.37 | 0.21 | 0.26 |
|  |  | 2 | 0.39 | 0.21 | 0.32 | 0.17 | 0.25 | 0.17 | 0.34 | 0.20 | 0.38 |
|  |  | 3 | 0.31 | 0.12 | 0.24 | 0.24 | 0.37 | 0.23 | 0.31 | 0.15 | 0.33 |
|  |  | 4 | 0.21 | 0.06 | 0.13 | 0.32 | 0.49 | 0.34 | 0.25 | 0.14 | 0.28 |
|  |  | 5 | -- | -- | -- | 0.41 | 0.59 | 0.44 | -- | -- | -- |
|  |  | >5 | -- | -- | -- | 0.12 | 0.27 | 0.21 | -- | -- | -- |
|  |  | Avg. | **0.35** | **0.18** | **0.22** | **0.23** | **0.37** | **0.26** | **0.35** | **0.20** | **0.32** |
| | | ($\geq 1$) | **0.35** | **0.19** | **0.25** | **0.23** | **0.37** | **0.26** | **0.32** | **0.17** | **0.31** |
|  | 30 | <1 | 0.36 | 0.18 | 0.11 | -- | -- | -- | 0.47 | 0.31 | 0.37 |
|  |  | 1 | 0.52 | 0.40 | 0.35 | 0.13 | 0.26 | 0.18 | 0.35 | 0.16 | 0.31 |
|  |  | 2 | 0.41 | 0.22 | 0.32 | 0.16 | 0.27 | 0.19 | 0.30 | 0.14 | 0.34 |
|  |  | 3 | 0.32 | 0.15 | 0.23 | 0.23 | 0.35 | 0.23 | 0.27 | 0.12 | 0.25 |
|  |  | 4 | 0.22 | 0.10 | 0.15 | 0.29 | 0.45 | 0.35 | 0.23 | 0.12 | 0.22 |
|  |  | 5 | -- | -- | -- | 0.35 | 0.51 | 0.46 | -- | -- | -- |
|  |  | >5 | -- | -- | -- | 0.07 | 0.17 | 0.21 | -- | -- | -- |
|  |  | Avg. | **0.37** | **0.21** | **0.23** | **0.20** | **0.34** | **0.27** | **0.33** | **0.17** | **0.29** |
| | | ($\geq 1$) | **0.37** | **0.22** | **0.26** | **0.20** | **0.34** | **0.27** | **0.29** | **0.13** | **0.28** |

**Web Table 4:**
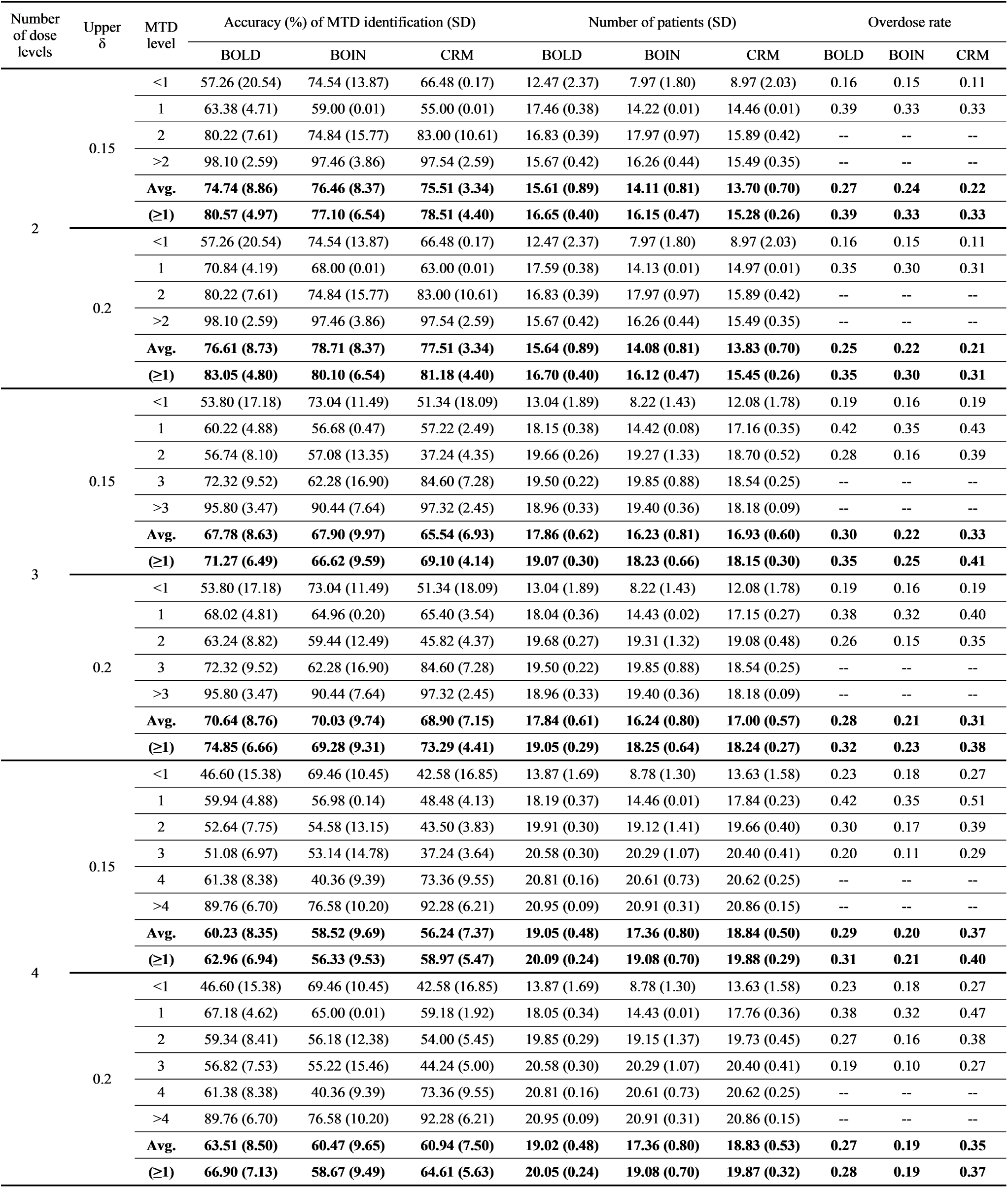
Summary of accuracy, efficiency, and overdosing rates of BOLD, BOIN and CRM by lattice (2, 3, 4-dose model) with non-informative prior for *φ* = 0.25 and *N_max_* = 21, utilizing random scenarios with upper *δ* = 0.15 and 0.2, where “Avg.” denotes the average for all the MTD levels and “(≥1)” denotes the average for the MTD levels that are ≥ 1.

| Number of dose levels | Upper $\delta$ | MTD level | Accuracy (%) of MTD identification (SD) | | | Number of patients (SD) | | | Overdose rate | | |
| --- | --- | --- | --- | --- | --- | --- | --- | --- | --- | --- | --- |
|  |  |  | BOLD | BOIN | CRM | BOLD | BOIN | CRM | BOLD | BOIN | CRM |
| 2 | 0.15 | <1 | 57.26 (20.54) | 74.54 (13.87) | 66.48 (0.17) | 12.47 (2.37) | 7.97 (1.80) | 8.97 (2.03) | 0.16 | 0.15 | 0.11 |
|  |  | 1 | 63.38 (4.71) | 59.00 (0.01) | 55.00 (0.01) | 17.46 (0.38) | 14.22 (0.01) | 14.46 (0.01) | 0.39 | 0.33 | 0.33 |
|  |  | 2 | 80.22 (7.61) | 74.84 (15.77) | 83.00 (10.61) | 16.83 (0.39) | 17.97 (0.97) | 15.89 (0.42) | -- | -- | -- |
|  |  | >2 | 98.10 (2.59) | 97.46 (3.86) | 97.54 (2.59) | 15.67 (0.42) | 16.26 (0.44) | 15.49 (0.35) | -- | -- | -- |
|  |  | **Avg.** | **74.74 (8.86)** | **76.46 (8.37)** | **75.51 (3.34)** | **15.61 (0.89)** | **14.11 (0.81)** | **13.70 (0.70)** | **0.27** | **0.24** | **0.22** |
|  |  | **($\geq 1$)** | **80.57 (4.97)** | **77.10 (6.54)** | **78.51 (4.40)** | **16.65 (0.40)** | **16.15 (0.47)** | **15.28 (0.26)** | **0.39** | **0.33** | **0.33** |
|  | 0.2 | <1 | 57.26 (20.54) | 74.54 (13.87) | 66.48 (0.17) | 12.47 (2.37) | 7.97 (1.80) | 8.97 (2.03) | 0.16 | 0.15 | 0.11 |
|  |  | 1 | 70.84 (4.19) | 68.00 (0.01) | 63.00 (0.01) | 17.59 (0.38) | 14.13 (0.01) | 14.97 (0.01) | 0.35 | 0.30 | 0.31 |
|  |  | 2 | 80.22 (7.61) | 74.84 (15.77) | 83.00 (10.61) | 16.83 (0.39) | 17.97 (0.97) | 15.89 (0.42) | -- | -- | -- |
|  |  | >2 | 98.10 (2.59) | 97.46 (3.86) | 97.54 (2.59) | 15.67 (0.42) | 16.26 (0.44) | 15.49 (0.35) | -- | -- | -- |
|  |  | **Avg.** | **76.61 (8.73)** | **78.71 (8.37)** | **77.51 (3.34)** | **15.64 (0.89)** | **14.08 (0.81)** | **13.83 (0.70)** | **0.25** | **0.22** | **0.21** |
|  |  | **($\geq 1$)** | **83.05 (4.80)** | **80.10 (6.54)** | **81.18 (4.40)** | **16.70 (0.40)** | **16.12 (0.47)** | **15.45 (0.26)** | **0.35** | **0.30** | **0.31** |
| 3 | 0.15 | <1 | 53.80 (17.18) | 73.04 (11.49) | 51.34 (18.09) | 13.04 (1.89) | 8.22 (1.43) | 12.08 (1.78) | 0.19 | 0.16 | 0.19 |
|  |  | 1 | 60.22 (4.88) | 56.68 (0.47) | 57.22 (2.49) | 18.15 (0.38) | 14.42 (0.08) | 17.16 (0.35) | 0.42 | 0.35 | 0.43 |
|  |  | 2 | 56.74 (8.10) | 57.08 (13.35) | 37.24 (4.35) | 19.66 (0.26) | 19.27 (1.33) | 18.70 (0.52) | 0.28 | 0.16 | 0.39 |
|  |  | 3 | 72.32 (9.52) | 62.28 (16.90) | 84.60 (7.28) | 19.50 (0.22) | 19.85 (0.88) | 18.54 (0.25) | -- | -- | -- |
|  |  | >3 | 95.80 (3.47) | 90.44 (7.64) | 97.32 (2.45) | 18.96 (0.33) | 19.40 (0.36) | 18.18 (0.09) | -- | -- | -- |
|  |  | **Avg.** | **67.78 (8.63)** | **67.90 (9.97)** | **65.54 (6.93)** | **17.86 (0.62)** | **16.23 (0.81)** | **16.93 (0.60)** | **0.30** | **0.22** | **0.33** |
|  |  | **($\geq 1$)** | **71.27 (6.49)** | **66.62 (9.59)** | **69.10 (4.14)** | **19.07 (0.30)** | **18.23 (0.66)** | **18.15 (0.30)** | **0.35** | **0.25** | **0.41** |
|  | 0.2 | <1 | 53.80 (17.18) | 73.04 (11.49) | 51.34 (18.09) | 13.04 (1.89) | 8.22 (1.43) | 12.08 (1.78) | 0.19 | 0.16 | 0.19 |
|  |  | 1 | 68.02 (4.81) | 64.96 (0.20) | 65.40 (3.54) | 18.04 (0.36) | 14.43 (0.02) | 17.15 (0.27) | 0.38 | 0.32 | 0.40 |
|  |  | 2 | 63.24 (8.82) | 59.44 (12.49) | 45.82 (4.37) | 19.68 (0.27) | 19.31 (1.32) | 19.08 (0.48) | 0.26 | 0.15 | 0.35 |
|  |  | 3 | 72.32 (9.52) | 62.28 (16.90) | 84.60 (7.28) | 19.50 (0.22) | 19.85 (0.88) | 18.54 (0.25) | -- | -- | -- |
|  |  | >3 | 95.80 (3.47) | 90.44 (7.64) | 97.32 (2.45) | 18.96 (0.33) | 19.40 (0.36) | 18.18 (0.09) | -- | -- | -- |
|  |  | **Avg.** | **70.64 (8.76)** | **70.03 (9.74)** | **68.90 (7.15)** | **17.84 (0.61)** | **16.24 (0.80)** | **17.00 (0.57)** | **0.28** | **0.21** | **0.31** |
|  |  | **($\geq 1$)** | **74.85 (6.66)** | **69.28 (9.31)** | **73.29 (4.41)** | **19.05 (0.29)** | **18.25 (0.64)** | **18.24 (0.27)** | **0.32** | **0.23** | **0.38** |
| 4 | 0.15 | <1 | 46.60 (15.38) | 69.46 (10.45) | 42.58 (16.85) | 13.87 (1.69) | 8.78 (1.30) | 13.63 (1.58) | 0.23 | 0.18 | 0.27 |
|  |  | 1 | 59.94 (4.88) | 56.98 (0.14) | 48.48 (4.13) | 18.19 (0.37) | 14.46 (0.01) | 17.84 (0.23) | 0.42 | 0.35 | 0.51 |
|  |  | 2 | 52.64 (7.75) | 54.58 (13.15) | 43.50 (3.83) | 19.91 (0.30) | 19.12 (1.41) | 19.66 (0.40) | 0.30 | 0.17 | 0.39 |
|  |  | 3 | 51.08 (6.97) | 53.14 (14.78) | 37.24 (3.64) | 20.58 (0.30) | 20.29 (1.07) | 20.40 (0.41) | 0.20 | 0.11 | 0.29 |
|  |  | 4 | 61.38 (8.38) | 40.36 (9.39) | 73.36 (9.55) | 20.81 (0.16) | 20.61 (0.73) | 20.62 (0.25) | -- | -- | -- |
|  |  | >4 | 89.76 (6.70) | 76.58 (10.20) | 92.28 (6.21) | 20.95 (0.09) | 20.91 (0.31) | 20.86 (0.15) | -- | -- | -- |
|  |  | **Avg.** | **60.23 (8.35)** | **58.52 (9.69)** | **56.24 (7.37)** | **19.05 (0.48)** | **17.36 (0.80)** | **18.84 (0.50)** | **0.29** | **0.20** | **0.37** |
|  |  | **($\geq 1$)** | **62.96 (6.94)** | **56.33 (9.53)** | **58.97 (5.47)** | **20.09 (0.24)** | **19.08 (0.70)** | **19.88 (0.29)** | **0.31** | **0.21** | **0.40** |
|  | 0.2 | <1 | 46.60 (15.38) | 69.46 (10.45) | 42.58 (16.85) | 13.87 (1.69) | 8.78 (1.30) | 13.63 (1.58) | 0.23 | 0.18 | 0.27 |
|  |  | 1 | 67.18 (4.62) | 65.00 (0.01) | 59.18 (1.92) | 18.05 (0.34) | 14.43 (0.01) | 17.76 (0.36) | 0.38 | 0.32 | 0.47 |
|  |  | 2 | 59.34 (8.41) | 56.18 (12.38) | 54.00 (5.45) | 19.85 (0.29) | 19.15 (1.37) | 19.73 (0.45) | 0.27 | 0.16 | 0.38 |
|  |  | 3 | 56.82 (7.53) | 55.22 (15.46) | 44.24 (5.00) | 20.58 (0.30) | 20.29 (1.07) | 20.40 (0.41) | 0.19 | 0.10 | 0.27 |
|  |  | 4 | 61.38 (8.38) | 40.36 (9.39) | 73.36 (9.55) | 20.81 (0.16) | 20.61 (0.73) | 20.62 (0.25) | -- | -- | -- |
|  |  | >4 | 89.76 (6.70) | 76.58 (10.20) | 92.28 (6.21) | 20.95 (0.09) | 20.91 (0.31) | 20.86 (0.15) | -- | -- | -- |
|  |  | **Avg.** | **63.51 (8.50)** | **60.47 (9.65)** | **60.94 (7.50)** | **19.02 (0.48)** | **17.36 (0.80)** | **18.83 (0.53)** | **0.27** | **0.19** | **0.35** |
|  |  | **($\geq 1$)** | **66.90 (7.13)** | **58.67 (9.49)** | **64.61 (5.63)** | **20.05 (0.24)** | **19.08 (0.70)** | **19.87 (0.32)** | **0.28** | **0.19** | **0.37** |

**Web Table 5:** Summary of accuracy, efficiency, and overdosing rates of BOLD, BOIN and CRM by lattice (2, 3, 4-dose model) with non-informative prior for *φ* = 0.25 and *N_max_* = 30, utilizing random scenarios with upper *δ* = 0.15 and 0.2, where “Avg.” denotes the average for all the MTD levels and “(≥1)” denotes the average for the MTD levels that are ≥ 1.

| Number of dose levels | Upper $\delta$ | MTD level | Accuracy (%) of MTD identification (SD) | | | Number of patients (SD) | | | Overdose rate | | |
| --- | --- | --- | --- | --- | --- | --- | --- | --- | --- | --- | --- |
|  |  |  | BOLD | BOIN | CRM | BOLD | BOIN | CRM | BOLD | BOIN | CRM |
| 2 | 0.15 | <1 | 57.52 (20.75) | 74.88 (14.53) | 56.44 (0.22) | 12.80 (2.53) | 8.17 (1.93) | 11.38 (2.18) | 0.15 | 0.15 | 0.17 |
|  |  | 1 | 62.34 (4.85) | 64.00 (0.01) | 52.00 (0.01) | 18.39 (0.46) | 16.29 (0.01) | 16.47 (0.01) | 0.37 | 0.34 | 0.42 |
|  |  | 2 | 82.82 (8.39) | 73.98 (12.66) | 89.34 (8.63) | 17.34 (0.54) | 20.29 (1.12) | 16.10 (0.32) | -- | -- | -- |
|  |  | >2 | 98.28 (2.37) | 97.84 (3.43) | 99.14 (1.50) | 15.70 (0.45) | 16.57 (0.63) | 15.35 (0.25) | -- | -- | -- |
|  |  | Avg. | **75.24 (9.09)** | **77.68 (7.66)** | **74.23 (2.59)** | **16.06 (0.99)** | **15.33 (0.92)** | **14.83 (0.69)** | **0.26** | **0.25** | **0.30** |
| | | ($\geq 1$) | **81.15 (5.20)** | **78.61 (5.37)** | **80.16 (3.38)** | **17.14 (0.48)** | **17.72 (0.58)** | **15.97 (0.19)** | **0.37** | **0.34** | **0.42** |
|  | 0.2 | <1 | 57.52 (20.75) | 74.88 (14.53) | 56.44 (8.02) | 12.80 (2.53) | 8.17 (1.93) | 11.38 (2.18) | 0.15 | 0.15 | 0.17 |
|  |  | 1 | 70.80 (4.34) | 70.00 (0.01) | 60.00 (0.01) | 18.69 (0.47) | 14.97 (0.01) | 16.50 (0.01) | 0.34 | 0.30 | 0.37 |
|  |  | 2 | 82.82 (8.39) | 73.98 (12.66) | 89.34 (8.63) | 17.34 (0.54) | 20.29 (1.12) | 16.10 (0.32) | -- | -- | -- |
|  |  | >2 | 98.28 (2.37) | 97.84 (3.43) | 99.14 (1.50) | 15.70 (0.45) | 16.57 (0.63) | 15.35 (0.25) | -- | -- | -- |
|  |  | Avg. | **77.36 (8.96)** | **79.18 (7.66)** | **76.23 (2.59)** | **16.13 (1.00)** | **15.00 (0.92)** | **14.83 (0.69)** | **0.25** | **0.22** | **0.27** |
| | | ($\geq 1$) | **83.97 (5.03)** | **80.61 (5.37)** | **82.83 (3.38)** | **17.25 (0.49)** | **17.28 (0.58)** | **15.98 (0.19)** | **0.34** | **0.30** | **0.37** |
| 3 | 0.15 | <1 | 54.62 (16.94) | 73.26 (12.11) | 53.16 (18.03) | 13.51 (2.11) | 8.50 (1.52) | 12.41 (2.02) | 0.19 | 0.16 | 0.18 |
|  |  | 1 | 61.66 (5.05) | 63.60 (0.67) | 59.90 (2.90) | 19.90 (0.55) | 17.02 (0.07) | 18.61 (0.23) | 0.43 | 0.37 | 0.42 |
|  |  | 2 | 59.06 (7.99) | 61.60 (11.26) | 43.12 (4.40) | 21.89 (0.61) | 24.11 (2.09) | 20.17 (0.93) | 0.29 | 0.17 | 0.36 |
|  |  | 3 | 74.68 (10.28) | 66.46 (15.65) | 85.26 (8.38) | 20.91 (0.54) | 24.08 (1.08) | 19.29 (0.42) | -- | -- | -- |
|  |  | >3 | 96.52 (3.44) | 91.90 (6.94) | 97.08 (2.69) | 19.39 (0.55) | 21.13 (1.01) | 18.51 (0.29) | -- | -- | -- |
|  |  | Avg. | **69.31 (8.74)** | **71.36 (9.33)** | **67.70 (7.28)** | **19.12 (0.87)** | **18.97 (1.15)** | **17.80 (0.78)** | **0.30** | **0.24** | **0.32** |
| | | ($\geq 1$) | **72.98 (6.69)** | **70.89 (8.63)** | **71.34 (4.59)** | **20.52 (0.56)** | **21.59 (1.06)** | **19.14 (0.47)** | **0.36** | **0.27** | **0.39** |
|  | 0.2 | <1 | 54.62 (16.94) | 73.26 (12.11) | 53.16 (18.03) | 13.51 (2.11) | 8.50 (1.52) | 12.41 (2.02) | 0.19 | 0.16 | 0.18 |
|  |  | 1 | 69.84 (4.31) | 66.96 (0.20) | 65.44 (1.16) | 19.69 (0.57) | 15.62 (0.06) | 18.12 (0.56) | 0.38 | 0.33 | 0.39 |
|  |  | 2 | 65.56 (7.96) | 65.10 (13.92) | 48.76 (6.37) | 21.80 (0.54) | 24.07 (2.02) | 20.72 (0.80) | 0.26 | 0.17 | 0.33 |
|  |  | 3 | 74.68 (10.28) | 66.46 (15.65) | 85.26 (8.38) | 20.91 (0.54) | 24.08 (1.08) | 19.29 (0.42) | -- | -- | -- |
|  |  | >3 | 96.52 (3.44) | 91.90 (6.94) | 97.08 (2.69) | 19.39 (0.55) | 21.13 (1.01) | 18.51 (0.29) | -- | -- | -- |
|  |  | Avg. | **72.24 (8.59)** | **72.74 (9.77)** | **69.94 (7.32)** | **19.06 (0.86)** | **18.68 (1.14)** | **17.81 (0.82)** | **0.28** | **0.22** | **0.30** |
| | | ($\geq 1$) | **76.65 (6.50)** | **72.61 (9.18)** | **74.14 (4.65)** | **20.45 (0.55)** | **21.23 (1.04)** | **19.16 (0.52)** | **0.32** | **0.25** | **0.36** |
| 4 | 0.15 | <1 | 47.08 (15.74) | 69.06 (11.48) | 43.10 (18.29) | 14.56 (1.99) | 9.22 (1.52) | 14.23 (1.88) | 0.24 | 0.19 | 0.27 |
|  |  | 1 | 61.14 (5.18) | 63.40 (0.76) | 50.14 (3.94) | 20.14 (0.49) | 17.20 (0.22) | 19.77 (0.41) | 0.44 | 0.38 | 0.52 |
|  |  | 2 | 57.76 (7.34) | 59.50 (13.89) | 50.90 (6.81) | 22.91 (0.54) | 24.19 (2.26) | 22.12 (0.92) | 0.33 | 0.20 | 0.38 |
|  |  | 3 | 53.70 (8.75) | 53.26 (13.43) | 44.26 (3.78) | 24.47 (0.45) | 26.99 (1.61) | 22.96 (0.54) | 0.23 | 0.13 | 0.28 |
|  |  | 4 | 67.94 (7.71) | 51.24 (12.52) | 74.48 (9.71) | 24.33 (0.43) | 26.72 (0.56) | 22.30 (0.37) | -- | -- | -- |
|  |  | >4 | 94.28 (4.32) | 88.44 (7.42) | 92.08 (5.57) | 23.21 (0.65) | 24.65 (0.59) | 21.85 (0.34) | -- | -- | -- |
|  |  | Avg. | **63.65 (8.17)** | **64.15 (9.92)** | **59.16 (8.02)** | **21.60 (0.76)** | **21.49 (1.13)** | **20.54 (0.74)** | **0.31** | **0.22** | **0.36** |
| | | ($\geq 1$) | **66.96 (6.66)** | **63.17 (9.60)** | **62.37 (5.57)** | **23.01 (0.51)** | **23.95 (1.05)** | **21.80 (0.52)** | **0.33** | **0.24** | **0.40** |
|  | 0.2 | <1 | 47.08 (15.74) | 69.06 (11.48) | 43.10 (18.29) | 14.56 (1.99) | 9.22 (1.52) | 14.23 (1.88) | 0.24 | 0.19 | 0.27 |
|  |  | 1 | 68.82 (4.48) | 67.52 (0.50) | 60.24 (3.85) | 19.71 (0.56) | 15.59 (0.05) | 18.62 (0.34) | 0.39 | 0.33 | 0.44 |
|  |  | 2 | 64.08 (7.75) | 62.94 (13.81) | 59.66 (5.29) | 22.52 (0.48) | 24.28 (2.11) | 21.83 (0.93) | 0.29 | 0.19 | 0.34 |
|  |  | 3 | 59.86 (9.11) | 58.58 (14.16) | 53.90 (5.44) | 24.57 (0.58) | 27.00 (1.46) | 23.58 (0.62) | 0.21 | 0.12 | 0.26 |
|  |  | 4 | 67.94 (7.71) | 51.24 (12.52) | 74.48 (9.71) | 24.33 (0.43) | 26.72 (0.56) | 22.30 (0.37) | -- | -- | -- |
|  |  | >4 | 94.28 (4.32) | 88.44 (7.42) | 92.08 (5.57) | 23.21 (0.65) | 24.65 (0.59) | 21.85 (0.34) | -- | -- | -- |
|  |  | Avg. | **67.01 (8.18)** | **66.30 (9.98)** | **63.91 (8.02)** | **21.48 (0.78)** | **21.24 (1.05)** | **20.40 (0.75)** | **0.28** | **0.21** | **0.33** |
| | | ($\geq 1$) | **71.00 (6.67)** | **65.74 (9.68)** | **68.07 (5.97)** | **22.87 (0.54)** | **23.65 (0.95)** | **21.64 (0.52)** | **0.30** | **0.21** | **0.35** |

**Web Table 6:** Summary of accuracy, efficiency and overdosing rates of BOLD, BOIN and CRM by PPAT threshold parameter *τ* at 5-dose model with non-informative prior for *φ* = 0.25 and 0.3 with *N_max_*= 21 and 30, assuming random scenarios of true DLT rates with upper *δ* = 0.15, where “Avg.” denotes the average for all the MTD levels and “(*≥*1)” denotes the average for the MTD levels that are *≥* 1.

| Target rate | N max | MTD level | Accuracy (%) of MTD identification (SD) |  |  | Number of patients (SD) |  |  | Overdose rate |  |  |
| --- | --- | --- | --- | --- | --- | --- | --- | --- | --- | --- | --- |
| | | | $\tau = 0.5$ (default) | $\tau = 0.48$ | $\tau = 0.45$ | $\tau = 0.5$ (default) | $\tau = 0.48$ | $\tau = 0.45$ | $\tau = 0.5$ (default) | $\tau = 0.48$ | $\tau = 0.45$ |
| 0.25 | 21 | <1 | 51.44 (14.09) | 51.72 (14.03) | 51.48 (14.31) | 13.39 (1.45) | 13.36 (1.48) | 13.24 (1.43) | 0.21 | 0.20 | 0.18 |
|  |  | 1 | 59.64 (4.51) | 61.42 (5.16) | 64.42 (5.19) | 18.24 (0.39) | 18.05 (0.34) | 17.75 (0.36) | 0.43 | 0.40 | 0.36 |
|  |  | 2 | 52.28 (7.32) | 52.22 (6.91) | 52.68 (8.87) | 19.92 (0.27) | 19.86 (0.22) | 19.62 (0.33) | 0.30 | 0.28 | 0.24 |
|  |  | 3 | 46.86 (8.94) | 45.64 (8.66) | 44.30 (8.11) | 20.56 (0.28) | 20.55 (0.31) | 20.29 (0.33) | 0.20 | 0.18 | 0.18 |
|  |  | 4 | 44.06 (7.74) | 42.36 (7.49) | 38.00 (7.11) | 20.81 (0.16) | 20.79 (0.19) | 20.63 (0.23) | 0.11 | 0.11 | 0.10 |
|  |  | 5 | 49.92 (8.67) | 49.08 (8.25) | 48.42 (8.26) | 20.91 (0.09) | 20.90 (0.09) | 20.80 (0.12) | -- | -- | -- |
|  |  | >5 | 78.50 (10.09) | 78.16 (10.13) | 77.42 (10.2) | 20.96 (0.05) | 20.96 (0.05) | 20.91 (0.09) | -- | -- | -- |
|  |  | **Avg.** | **54.67 (8.77)** | **54.37 (8.66)** | **53.82 (8.86)** | **19.26 (0.38)** | **19.21 (0.38)** | **19.04 (0.41)** | **0.25** | **0.23** | **0.21** |
|  |  | **($\geq 1$)** | **55.21 (8.00)** | **54.75 (7.90)** | **54.15 (8.08)** | **20.23 (0.21)** | **20.19 (0.20)** | **20.00 (0.24)** | **0.26** | **0.24** | **0.22** |
|  | 30 | <1 | 52.26 (13.81) | 52.30 (13.60) | 52.46 (13.78) | 14.07 (1.71) | 13.93 (1.71) | 13.72 (1.63) | 0.23 | 0.21 | 0.19 |
|  |  | 1 | 60.34 (5.17) | 62.36 (5.10) | 64.78 (5.75) | 20.21 (0.51) | 19.67 (0.43) | 19.08 (0.57) | 0.44 | 0.41 | 0.37 |
|  |  | 2 | 55.64 (6.76) | 54.46 (8.70) | 58.84 (9.72) | 23.07 (0.51) | 22.76 (0.53) | 22.13 (0.54) | 0.33 | 0.31 | 0.27 |
|  |  | 3 | 51.00 (9.18) | 49.98 (8.70) | 45.42 (8.96) | 25.24 (0.51) | 25.02 (0.51) | 23.95 (0.62) | 0.26 | 0.24 | 0.21 |
|  |  | 4 | 47.70 (7.01) | 46.74 (6.92) | 41.32 (7.92) | 26.95 (0.46) | 26.71 (0.47) | 25.53 (0.49) | 0.18 | 0.16 | 0.14 |
|  |  | 5 | 63.40 (8.66) | 60.76 (9.06) | 54.56 (9.03) | 26.90 (0.44) | 26.77 (0.39) | 26.00 (0.49) | -- | -- | -- |
|  |  | >5 | 90.76 (5.02) | 89.04 (5.64) | 83.00 (8.74) | 26.36 (0.62) | 26.33 (0.60) | 25.90 (0.39) | -- | -- | -- |
|  |  | **Avg.** | **60.16 (7.95)** | **59.38 (8.25)** | **57.20 (9.13)** | **23.26 (0.68)** | **23.03 (0.66)** | **22.33 (0.68)** | **0.29** | **0.27** | **0.24** |
|  |  | **($\geq 1$)** | **61.47 (7.11)** | **60.56 (7.48)** | **57.99 (8.46)** | **24.57 (0.53)** | **24.33 (0.51)** | **23.56 (0.54)** | **0.30** | **0.28** | **0.25** |
| 0.3 | 21 | <1 | 49.86 (11.18) | 55.10 (9.66) | 55.82 (10.01) | 13.78 (1.38) | 12.05 (1.13) | 11.96 (1.16) | 0.36 | 0.18 | 0.15 |
|  |  | 1 | 50.76 (5.97) | 55.74 (4.93) | 60.90 (5.54) | 18.16 (0.44) | 16.99 (0.46) | 17.17 (0.51) | 0.51 | 0.38 | 0.33 |
|  |  | 2 | 49.16 (8.79) | 51.24 (7.64) | 54.34 (9.48) | 19.82 (0.33) | 19.25 (0.41) | 19.69 (0.41) | 0.39 | 0.25 | 0.20 |
|  |  | 3 | 44.50 (7.04) | 46.38 (9.41) | 45.54 (10.52) | 20.47 (0.28) | 20.08 (0.45) | 20.38 (0.40) | 0.31 | 0.17 | 0.13 |
|  |  | 4 | 43.48 (7.53) | 42.46 (7.63) | 38.48 (7.89) | 20.78 (0.16) | 20.54 (0.25) | 20.69 (0.21) | 0.21 | 0.08 | 0.07 |
|  |  | 5 | 58.72 (7.75) | 47.16 (11.62) | 42.34 (10.89) | 20.87 (0.13) | 20.74 (0.20) | 20.83 (0.16) | -- | -- | -- |
|  |  | >5 | 87.76 (7.32) | 77.02 (12.2) | 72.94 (12.98) | 20.94 (0.08) | 20.92 (0.09) | 20.94 (0.07) | -- | -- | -- |
|  |  | **Avg.** | **54.89 (7.94)** | **53.59 (9.01)** | **52.91 (9.62)** | **19.26 (0.4)** | **18.65 (0.43)** | **18.81 (0.42)** | **0.35** | **0.21** | **0.18** |
|  |  | **($\geq 1$)** | **55.73 (7.48)** | **53.37 (8.92)** | **52.49 (9.56)** | **20.17 (0.24)** | **19.75 (0.31)** | **19.95 (0.29)** | **0.35** | **0.22** | **0.18** |
|  | 30 | <1 | 53.20 (10.89) | 55.62 (9.86) | 55.66 (10.21) | 14.98 (1.66) | 12.45 (1.32) | 12.31 (1.33) | 0.36 | 0.19 | 0.15 |
|  |  | 1 | 51.86 (4.42) | 57.06 (5.95) | 63.34 (5.60) | 20.77 (0.71) | 18.51 (0.60) | 18.45 (0.60) | 0.52 | 0.40 | 0.35 |
|  |  | 2 | 54.10 (7.08) | 57.78 (7.94) | 59.42 (10.78) | 23.49 (0.56) | 21.81 (0.59) | 22.33 (0.60) | 0.41 | 0.28 | 0.23 |
|  |  | 3 | 49.72 (9.18) | 48.88 (8.85) | 47.70 (10.57) | 24.94 (0.70) | 24.15 (0.70) | 24.31 (0.67) | 0.32 | 0.21 | 0.17 |
|  |  | 4 | 48.20 (8.18) | 42.68 (8.36) | 39.12 (8.64) | 26.02 (0.50) | 25.85 (0.58) | 25.98 (0.46) | 0.22 | 0.15 | 0.11 |
|  |  | 5 | 65.36 (7.95) | 58.64 (10.8) | 51.70 (11.25) | 25.51 (0.43) | 26.46 (0.55) | 26.77 (0.57) | -- | -- | -- |
|  |  | >5 | 92.84 (5.20) | 89.64 (6.15) | 84.74 (8.74) | 24.63 (0.32) | 26.40 (0.54) | 26.48 (0.52) | -- | -- | -- |
|  |  | **Avg.** | **59.33 (7.56)** | **58.61 (8.27)** | **57.38 (9.40)** | **22.90 (0.70)** | **22.23 (0.70)** | **22.37 (0.68)** | **0.37** | **0.25** | **0.20** |
|  |  | **($\geq 1$)** | **60.35 (7.08)** | **59.11 (8.05)** | **57.67 (9.28)** | **24.23 (0.54)** | **23.86 (0.59)** | **24.05 (0.57)** | **0.37** | **0.26** | **0.22** |

**Web Table 7:**
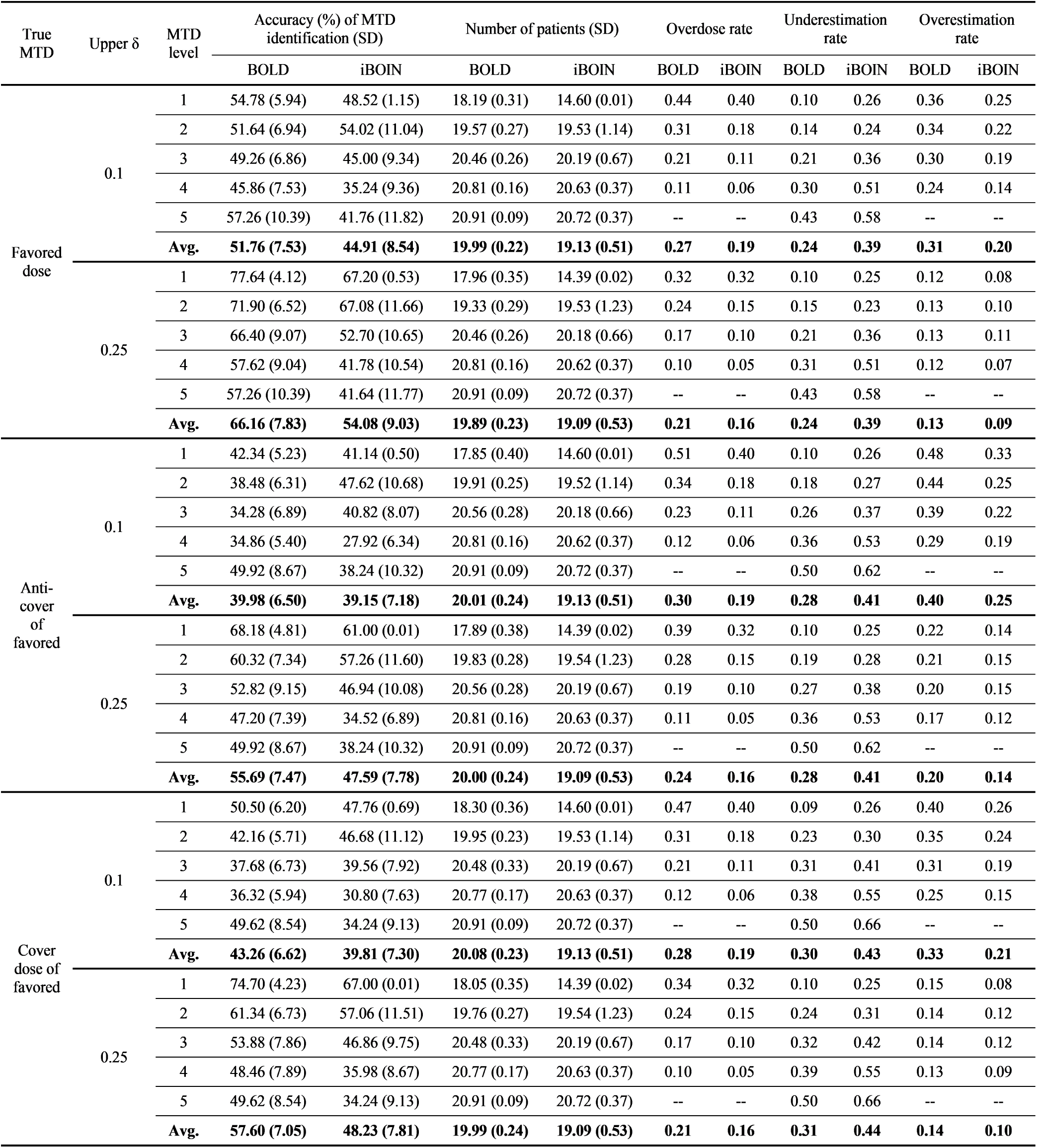
Summary of accuracy, efficiency, overdose, underestimation, and overestimation rates of BOLD and iBOIN, utilizing prior PESS specification with 5-dose lattice, *φ* = 0.25, *N_max_* = 21, and chosen upper *δ* values of 0.1 and 0.25 with random scenarios.

| True MTD | Upper $\delta$ | MTD level | Accuracy (%) of MTD identification (SD) | | Number of patients (SD) | | Overdose rate | | Underestimation rate | | Overestimation rate | |
| --- | --- | --- | --- | --- | --- | --- | --- | --- | --- | --- | --- | --- |
|  |  |  | BOLD | iBOIN | BOLD | iBOIN | BOLD | iBOIN | BOLD | iBOIN | BOLD | iBOIN |
| Favored dose | 0.1 | 1 | 54.78 (5.94) | 48.52 (1.15) | 18.19 (0.31) | 14.60 (0.01) | 0.44 | 0.40 | 0.10 | 0.26 | 0.36 | 0.25 |
|  |  | 2 | 51.64 (6.94) | 54.02 (11.04) | 19.57 (0.27) | 19.53 (1.14) | 0.31 | 0.18 | 0.14 | 0.24 | 0.34 | 0.22 |
|  |  | 3 | 49.26 (6.86) | 45.00 (9.34) | 20.46 (0.26) | 20.19 (0.67) | 0.21 | 0.11 | 0.21 | 0.36 | 0.30 | 0.19 |
|  |  | 4 | 45.86 (7.53) | 35.24 (9.36) | 20.81 (0.16) | 20.63 (0.37) | 0.11 | 0.06 | 0.30 | 0.51 | 0.24 | 0.14 |
|  |  | 5 | 57.26 (10.39) | 41.76 (11.82) | 20.91 (0.09) | 20.72 (0.37) | -- | -- | 0.43 | 0.58 | -- | -- |
|  |  | Avg. | **51.76 (7.53)** | **44.91 (8.54)** | **19.99 (0.22)** | **19.13 (0.51)** | **0.27** | **0.19** | **0.24** | **0.39** | **0.31** | **0.20** |
|  | 0.25 | 1 | 77.64 (4.12) | 67.20 (0.53) | 17.96 (0.35) | 14.39 (0.02) | 0.32 | 0.32 | 0.10 | 0.25 | 0.12 | 0.08 |
|  |  | 2 | 71.90 (6.52) | 67.08 (11.66) | 19.33 (0.29) | 19.53 (1.23) | 0.24 | 0.15 | 0.15 | 0.23 | 0.13 | 0.10 |
|  |  | 3 | 66.40 (9.07) | 52.70 (10.65) | 20.46 (0.26) | 20.18 (0.66) | 0.17 | 0.10 | 0.21 | 0.36 | 0.13 | 0.11 |
|  |  | 4 | 57.62 (9.04) | 41.78 (10.54) | 20.81 (0.16) | 20.62 (0.37) | 0.10 | 0.05 | 0.31 | 0.51 | 0.12 | 0.07 |
|  |  | 5 | 57.26 (10.39) | 41.64 (11.77) | 20.91 (0.09) | 20.72 (0.37) | -- | -- | 0.43 | 0.58 | -- | -- |
|  |  | Avg. | **66.16 (7.83)** | **54.08 (9.03)** | **19.89 (0.23)** | **19.09 (0.53)** | **0.21** | **0.16** | **0.24** | **0.39** | **0.13** | **0.09** |
| Anti-cover of favored | 0.1 | 1 | 42.34 (5.23) | 41.14 (0.50) | 17.85 (0.40) | 14.60 (0.01) | 0.51 | 0.40 | 0.10 | 0.26 | 0.48 | 0.33 |
|  |  | 2 | 38.48 (6.31) | 47.62 (10.68) | 19.91 (0.25) | 19.52 (1.14) | 0.34 | 0.18 | 0.18 | 0.27 | 0.44 | 0.25 |
|  |  | 3 | 34.28 (6.89) | 40.82 (8.07) | 20.56 (0.28) | 20.18 (0.66) | 0.23 | 0.11 | 0.26 | 0.37 | 0.39 | 0.22 |
|  |  | 4 | 34.86 (5.40) | 27.92 (6.34) | 20.81 (0.16) | 20.62 (0.37) | 0.12 | 0.06 | 0.36 | 0.53 | 0.29 | 0.19 |
|  |  | 5 | 49.92 (8.67) | 38.24 (10.32) | 20.91 (0.09) | 20.72 (0.37) | -- | -- | 0.50 | 0.62 | -- | -- |
|  |  | Avg. | **39.98 (6.50)** | **39.15 (7.18)** | **20.01 (0.24)** | **19.13 (0.51)** | **0.30** | **0.19** | **0.28** | **0.41** | **0.40** | **0.25** |
|  | 0.25 | 1 | 68.18 (4.81) | 61.00 (0.01) | 17.89 (0.38) | 14.39 (0.02) | 0.39 | 0.32 | 0.10 | 0.25 | 0.22 | 0.14 |
|  |  | 2 | 60.32 (7.34) | 57.26 (11.60) | 19.83 (0.28) | 19.54 (1.23) | 0.28 | 0.15 | 0.19 | 0.28 | 0.21 | 0.15 |
|  |  | 3 | 52.82 (9.15) | 46.94 (10.08) | 20.56 (0.28) | 20.19 (0.67) | 0.19 | 0.10 | 0.27 | 0.38 | 0.20 | 0.15 |
|  |  | 4 | 47.20 (7.39) | 34.52 (6.89) | 20.81 (0.16) | 20.63 (0.37) | 0.11 | 0.05 | 0.36 | 0.53 | 0.17 | 0.12 |
|  |  | 5 | 49.92 (8.67) | 38.24 (10.32) | 20.91 (0.09) | 20.72 (0.37) | -- | -- | 0.50 | 0.62 | -- | -- |
|  |  | Avg. | **55.69 (7.47)** | **47.59 (7.78)** | **20.00 (0.24)** | **19.09 (0.53)** | **0.24** | **0.16** | **0.28** | **0.41** | **0.20** | **0.14** |
| Cover dose of favored | 0.1 | 1 | 50.50 (6.20) | 47.76 (0.69) | 18.30 (0.36) | 14.60 (0.01) | 0.47 | 0.40 | 0.09 | 0.26 | 0.40 | 0.26 |
|  |  | 2 | 42.16 (5.71) | 46.68 (11.12) | 19.95 (0.23) | 19.53 (1.14) | 0.31 | 0.18 | 0.23 | 0.30 | 0.35 | 0.24 |
|  |  | 3 | 37.68 (6.73) | 39.56 (7.92) | 20.48 (0.33) | 20.19 (0.67) | 0.21 | 0.11 | 0.31 | 0.41 | 0.31 | 0.19 |
|  |  | 4 | 36.32 (5.94) | 30.80 (7.63) | 20.77 (0.17) | 20.63 (0.37) | 0.12 | 0.06 | 0.38 | 0.55 | 0.25 | 0.15 |
|  |  | 5 | 49.62 (8.54) | 34.24 (9.13) | 20.91 (0.09) | 20.72 (0.37) | -- | -- | 0.50 | 0.66 | -- | -- |
|  |  | Avg. | **43.26 (6.62)** | **39.81 (7.30)** | **20.08 (0.23)** | **19.13 (0.51)** | **0.28** | **0.19** | **0.30** | **0.43** | **0.33** | **0.21** |
|  | 0.25 | 1 | 74.70 (4.23) | 67.00 (0.01) | 18.05 (0.35) | 14.39 (0.02) | 0.34 | 0.32 | 0.10 | 0.25 | 0.15 | 0.08 |
|  |  | 2 | 61.34 (6.73) | 57.06 (11.51) | 19.76 (0.27) | 19.54 (1.23) | 0.24 | 0.15 | 0.24 | 0.31 | 0.14 | 0.12 |
|  |  | 3 | 53.88 (7.86) | 46.86 (9.75) | 20.48 (0.33) | 20.19 (0.67) | 0.17 | 0.10 | 0.32 | 0.42 | 0.14 | 0.12 |
|  |  | 4 | 48.46 (7.89) | 35.98 (8.67) | 20.77 (0.17) | 20.63 (0.37) | 0.10 | 0.05 | 0.39 | 0.55 | 0.13 | 0.09 |
|  |  | 5 | 49.62 (8.54) | 34.24 (9.13) | 20.91 (0.09) | 20.72 (0.37) | -- | -- | 0.50 | 0.66 | -- | -- |
|  |  | Avg. | **57.60 (7.05)** | **48.23 (7.81)** | **19.99 (0.24)** | **19.09 (0.53)** | **0.21** | **0.16** | **0.31** | **0.44** | **0.14** | **0.10** |

**Web Table 8:**
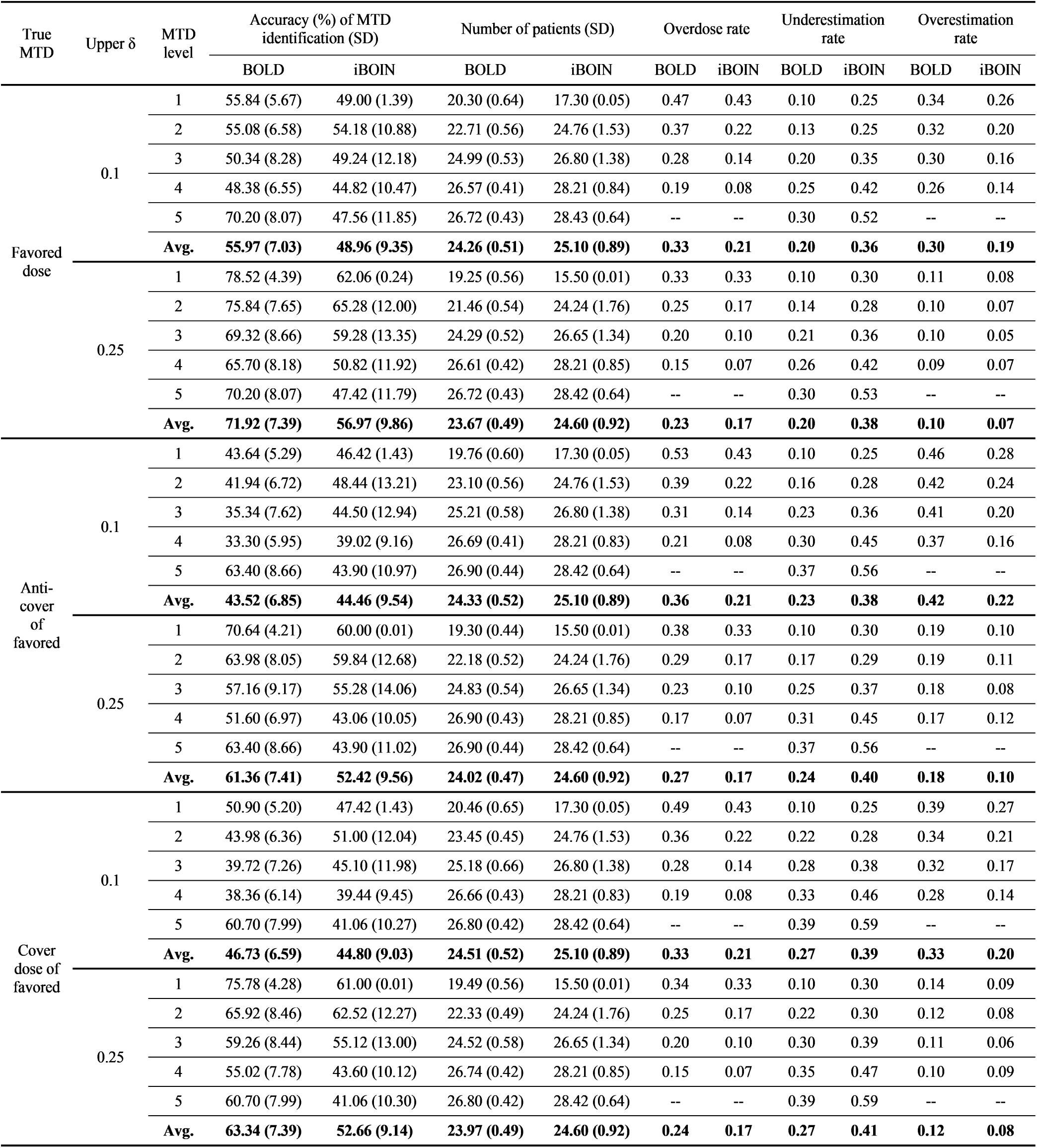
Summary of accuracy, efficiency, overdose, underestimation, and overestimation rates of BOLD and iBOIN, utilizing prior PESS specification with 5-dose lattice, *φ* = 0.25, *N_max_* = 30, and chosen upper *δ* values of 0.1 and 0.25 with random scenarios.

**Web Table 9:**
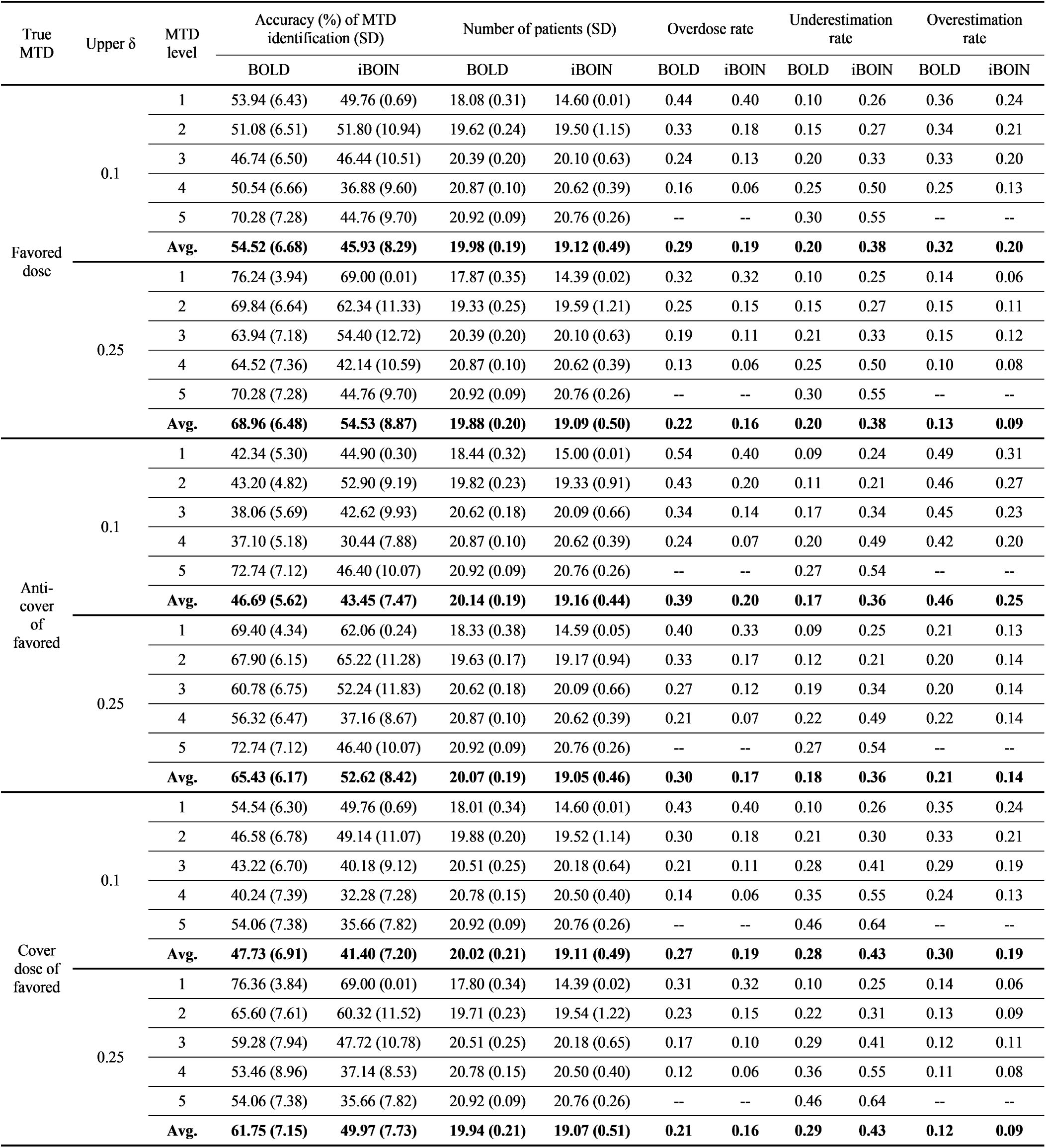
Summary of accuracy, efficiency, overdose, underestimation, and overestimation rates of BOLD and iBOIN, utilizing prior mean/skeleton specification with 5-dose lattice, *φ* = 0.25, *N_max_* = 21, and chosen upper *δ* values of 0.1 and 0.25 with random scenarios.

**Web Table 10:** Summary of accuracy, efficiency, overdose, underestimation, and overestimation rates of BOLD and iBOIN, utilizing prior mean/skeleton specification with 5-dose lattice, *φ* = 0.25, *N_max_* = 30, and chosen upper *δ* values of 0.1 and 0.25 with random scenarios.

| True MTD | Upper $\delta$ | MTD level | Accuracy (%) of MTD identification (SD) | | Number of patients (SD) | | Overdose rate | | Underestimation rate | | Overestimation rate | |
| --- | --- | --- | --- | --- | --- | --- | --- | --- | --- | --- | --- | --- |
|  |  |  | BOLD | iBOIN | BOLD | iBOIN | BOLD | iBOIN | BOLD | iBOIN | BOLD | iBOIN |
| Favored dose | 0.1 | 1 | 54.52 (5.28) | 49.00 (1.39) | 19.98 (0.62) | 17.30 (0.05) | 0.46 | 0.43 | 0.10 | 0.25 | 0.35 | 0.26 |
|  |  | 2 | 52.40 (7.06) | 52.30 (12.25) | 22.40 (0.48) | 24.87 (1.36) | 0.37 | 0.22 | 0.14 | 0.26 | 0.34 | 0.21 |
|  |  | 3 | 49.04 (7.18) | 48.88 (12.31) | 24.33 (0.51) | 26.82 (1.40) | 0.30 | 0.16 | 0.19 | 0.33 | 0.32 | 0.18 |
|  |  | 4 | 47.84 (5.22) | 42.80 (9.00) | 25.77 (0.33) | 27.71 (0.84) | 0.22 | 0.10 | 0.22 | 0.43 | 0.30 | 0.14 |
|  |  | 5 | 73.08 (6.68) | 51.82 (10.42) | 25.20 (0.30) | 27.92 (0.49) | -- | -- | 0.27 | 0.48 | -- | -- |
|  |  | Avg. | **55.38 (6.28)** | **48.96 (9.07)** | **23.54 (0.45)** | **24.92 (0.83)** | **0.34** | **0.23** | **0.18** | **0.35** | **0.33** | **0.20** |
|  | 0.25 | 1 | 77.02 (4.61) | 63.00 (0.01) | 19.06 (0.50) | 15.50 (0.01) | 0.32 | 0.33 | 0.10 | 0.30 | 0.13 | 0.07 |
|  |  | 2 | 73.40 (7.19) | 64.44 (11.98) | 21.28 (0.54) | 24.54 (1.55) | 0.26 | 0.17 | 0.15 | 0.28 | 0.11 | 0.08 |
|  |  | 3 | 69.38 (6.48) | 61.64 (13.30) | 23.64 (0.47) | 26.68 (1.36) | 0.21 | 0.12 | 0.20 | 0.33 | 0.10 | 0.05 |
|  |  | 4 | 66.94 (6.82) | 53.56 (10.34) | 25.80 (0.32) | 27.97 (1.00) | 0.16 | 0.09 | 0.24 | 0.41 | 0.09 | 0.05 |
|  |  | 5 | 73.08 (6.68) | 51.82 (10.45) | 25.20 (0.30) | 27.91 (0.49) | -- | -- | 0.27 | 0.48 | -- | -- |
|  |  | Avg. | **71.96 (6.36)** | **58.89 (9.21)** | **22.99 (0.43)** | **24.52 (0.88)** | **0.24** | **0.18** | **0.19** | **0.36** | **0.11** | **0.06** |
| Anti-cover of favored | 0.1 | 1 | 43.48 (4.42) | 46.66 (0.77) | 20.24 (0.63) | 17.41 (0.05) | 0.55 | 0.43 | 0.09 | 0.27 | 0.47 | 0.26 |
|  |  | 2 | 44.72 (5.53) | 53.98 (8.95) | 22.60 (0.46) | 24.47 (1.48) | 0.46 | 0.25 | 0.11 | 0.22 | 0.45 | 0.24 |
|  |  | 3 | 41.02 (5.57) | 49.20 (10.46) | 24.56 (0.48) | 26.48 (1.11) | 0.39 | 0.18 | 0.15 | 0.31 | 0.44 | 0.20 |
|  |  | 4 | 40.96 (5.39) | 42.22 (8.05) | 25.62 (0.33) | 27.95 (0.84) | 0.29 | 0.12 | 0.18 | 0.38 | 0.41 | 0.19 |
|  |  | 5 | 77.26 (6.52) | 52.74 (10.67) | 25.09 (0.28) | 27.54 (0.62) | -- | -- | 0.23 | 0.47 | -- | -- |
|  |  | Avg. | **49.49 (5.49)** | **48.96 (7.78)** | **23.62 (0.44)** | **24.77 (0.82)** | **0.42** | **0.24** | **0.15** | **0.33** | **0.44** | **0.22** |
|  | 0.25 | 1 | 69.98 (3.80) | 61.00 (0.01) | 19.55 (0.48) | 15.50 (0.01) | 0.39 | 0.33 | 0.10 | 0.30 | 0.20 | 0.09 |
|  |  | 2 | 70.00 (6.71) | 69.48 (10.77) | 21.75 (0.53) | 23.90 (1.19) | 0.33 | 0.18 | 0.11 | 0.22 | 0.19 | 0.09 |
|  |  | 3 | 66.08 (6.32) | 59.28 (12.19) | 24.26 (0.37) | 26.03 (1.37) | 0.28 | 0.13 | 0.16 | 0.31 | 0.18 | 0.10 |
|  |  | 4 | 62.34 (5.97) | 51.22 (7.15) | 26.22 (0.33) | 27.73 (0.81) | 0.23 | 0.09 | 0.21 | 0.40 | 0.17 | 0.09 |
|  |  | 5 | 77.26 (6.52) | 52.74 (10.67) | 25.09 (0.28) | 27.54 (0.62) | -- | -- | 0.23 | 0.47 | -- | -- |
|  |  | Avg. | **69.13 (5.86)** | **58.74 (8.16)** | **23.37 (0.40)** | **24.14 (0.80)** | **0.31** | **0.18** | **0.16** | **0.34** | **0.18** | **0.09** |
| Cover dose of favored | 0.1 | 1 | 55.10 (5.59) | 49.00 (1.39) | 19.73 (0.60) | 17.30 (0.05) | 0.45 | 0.43 | 0.10 | 0.25 | 0.34 | 0.26 |
|  |  | 2 | 48.42 (7.42) | 52.20 (12.12) | 23.02 (0.54) | 24.76 (1.53) | 0.33 | 0.22 | 0.20 | 0.28 | 0.31 | 0.20 |
|  |  | 3 | 43.96 (7.30) | 48.02 (11.57) | 24.77 (0.61) | 26.86 (1.26) | 0.28 | 0.13 | 0.26 | 0.37 | 0.30 | 0.15 |
|  |  | 4 | 42.24 (6.21) | 41.56 (9.68) | 26.07 (0.36) | 27.94 (0.96) | 0.20 | 0.10 | 0.31 | 0.45 | 0.27 | 0.14 |
|  |  | 5 | 64.82 (7.88) | 43.82 (9.52) | 25.80 (0.30) | 28.11 (0.60) | -- | -- | 0.35 | 0.56 | -- | -- |
|  |  | Avg. | **50.91 (6.88)** | **46.92 (8.86)** | **23.88 (0.48)** | **24.99 (0.88)** | **0.31** | **0.22** | **0.25** | **0.38** | **0.31** | **0.19** |
|  | 0.25 | 1 | 77.52 (4.59) | 63.00 (0.01) | 18.99 (0.52) | 15.50 (0.01) | 0.31 | 0.33 | 0.10 | 0.30 | 0.12 | 0.07 |
|  |  | 2 | 68.94 (8.97) | 64.14 (11.92) | 22.01 (0.51) | 24.24 (1.76) | 0.24 | 0.17 | 0.20 | 0.30 | 0.11 | 0.06 |
|  |  | 3 | 62.54 (9.07) | 57.78 (14.49) | 24.19 (0.50) | 26.73 (1.21) | 0.19 | 0.10 | 0.27 | 0.38 | 0.10 | 0.04 |
|  |  | 4 | 58.78 (7.66) | 48.36 (8.91) | 26.13 (0.45) | 28.07 (0.99) | 0.15 | 0.08 | 0.32 | 0.46 | 0.09 | 0.05 |
|  |  | 5 | 64.82 (7.88) | 43.82 (9.52) | 25.80 (0.30) | 28.11 (0.60) | -- | -- | 0.35 | 0.56 | -- | -- |
|  |  | Avg. | **66.52 (7.63)** | **55.42 (8.97)** | **23.43 (0.45)** | **24.53 (0.91)** | **0.22** | **0.17** | **0.25** | **0.40** | **0.10** | **0.06** |

